# Ischemia promotes hypertrophic nerve trunk formation and enteric neuron cell death in Hirschsprung’s disease

**DOI:** 10.1101/2024.03.14.24304192

**Authors:** Deshu Xu, Weiwei Liang, Chaoting Lan, Lanying Li, Weiyong Zhong, Meng Yao, Xiaoyu Zuo, Jixiao Zeng, Wei Zhong, Qiang Wu, Andrew M Lew, Wenhao Zhou, Huimin Xia, Fan Bai, Yuxia Zhang, Yan Zhang

## Abstract

**Objective:** Enteric nervous system dysfunction is linked to digestive and neurological disorders. Hirschsprung’s disease (HSCR) is characterized by the loss of enteric neuron cells (ENCs) in the distal colon. Embryonic enteric neural crest cell (ENCC) migration defects contribute to HSCR development in some infants, but postnatal factors that regulate ENC fate are undetermined. We sought to establish how postnatal changes contribute to HSCR by profiling the colonic microenvironment of HSCR infants.

**Design:** In this study, we recruited infants with HSCR, infants with anorectal malformations but normal ENC development (CT), and a group of age-matched healthy control subjects. Laboratory findings and clinical manifestations were recorded. Single-cell and spatial transcriptome sequencing were applied to colonic tissues from a sub-cohort of CT and HSCR infants. Patient specimens, a mouse model of neonatal ischemic enterocolitis, and *Sox10* knockdown mouse (*Sox10*^WT/MUT^) were used to reveal the factors that lead to ENC loss in HSCR infants.

**Results:** We discover that intestinal ischemia promotes CLDN1^+^ hypertrophic nerve trunk formation and ENC death. Mechanistically, ischemia leads to defective nitric oxide (NO) signaling in ENCs, which aggravates mitochondria damage and caspase-mediated apoptosis that can be ameliorated by a NO donor drug.

**Conclusion:** We show that ischemia contributes to postnatal ENC loss in HSCR infants and suggest that NO donor drugs may alleviate ischemia-related ENC death.

## WHAT IS ALREADY KNOWN ON THIS TOPIC

- Hirschsprung’s disease (HSCR) is characterized by the absence of enteric nerve cells (ENCs) in distal colons, which is caused by defective enteric neural crest cell (ENCC) migration and colonization during the embryonic stage. However, extensive ENCC-derived glial cells can be observed in aganglionic colons of HSCR patients.

## WHAT THIS STUDY ADDS

- HSCR patients display ischemia-like signatures in the colon, highlighted by extensive platelet-collagen aggregation in the blood vessels of both ganglionic and aganglionic colon biopsies.
- In ganglionic colon biopsies of HSCR infants, ENCs present ischemic phenotypes, including zinc ion accumulation, mitochondria damage, and Caspase-mediated apoptosis. A nitric oxide donor drug ameliorates ischemia-induced ENC death by promoting mitophagy.
- Hypertrophic nerve trunks (HNTs) are wrapped by *CLDN1*^+^ fibroblasts (*CLDN1*^+^FLCs). This structure is formed by ischemic stress, a neuron-derived growth factor (FGF1), and an inflammatory signal (TNF-α).

## HOW THIS STUDY MIGHT AFFECT RESEARCH, PRACTICE OR POLICY

- Our results reveal that ischemia-induced postnatal ENC loss is a novel pathogenic mechanism of HSCR and suggest that NO donor drugs may be useful in treating ischemia-related ENC death.

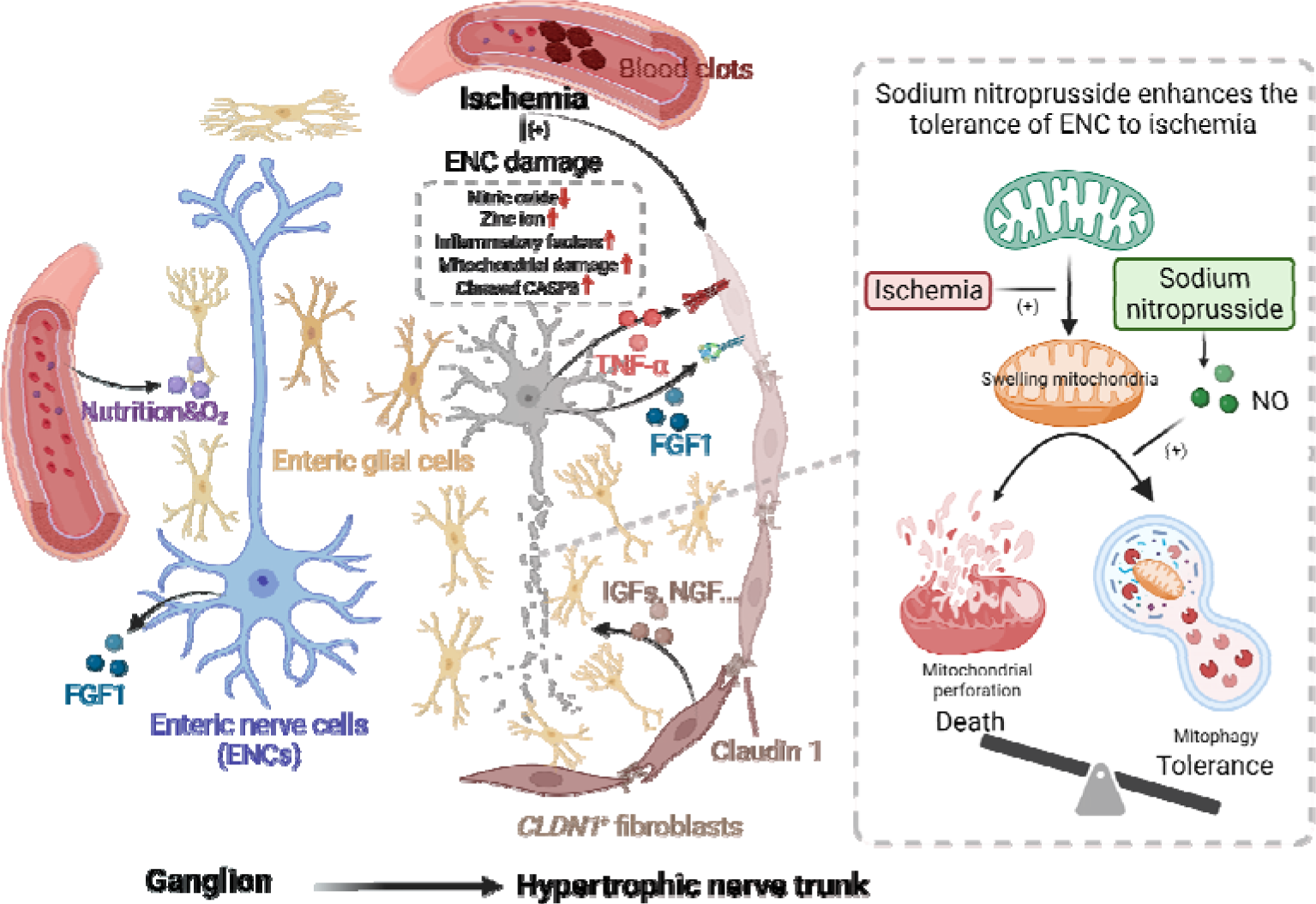
The schematic finding of the study. The intestines of infants with HSCR display extensive ischemia, highlighted by platelet-collagen aggregation in the blood vessels. *CLDN1*^+^ fibroblasts are differentiated under ischemia stress with a neuron-derived growth factor (FGF1) and an inflammatory cytokine (TNF-α). Nitric oxide (NO) synthesis is reduced in enteric nerve cells (ENCs) under ischemia, leading to zinc accumulation, mitochondria damage, and caspase-dependent cell death. Providing an NO donor drug (sodium nitroprusside) prevented ENC death by inducing mitophagy in a neonatal ischemic enterocolitis (NIE) model.

## INTRODUCTION

The enteric nervous system (ENS) is the largest component of the autonomic nervous system and orchestrates gastrointestinal homeostasis and motility. ENS dysfunction is linked to a vast range of digestive diseases and neurological disorders including inflammatory bowel disease and autism^1^ ^2^. During embryonic stage, the enteric neural crest cells (ENCCs) migrate from the foregut toward the distal hindgut, colonize, and differentiate into enteric nerve cells (ENCs) and glial cells^3^. Defective ENCC migration is linked to Hirschsprung’s disease (HSCR), an intestinal motility disorder leading to severe intestinal obstruction in infancy. The disease is characterized by the absence of ENCs in the distal colon^4^. Previous studies have shown that genetic predispositions (such as variants of *RET*, *SOX10*, and *EDNRB*) and nutritional factors (such as retinoic acid deficiency) contribute to HSCR by impairing ENCC migration^4–10^. However, HSCR does not follow simple mendelian inheritance^5^ ^10^, and extensive ENCC-derived enteric glial cells have been observed in aganglionic colons in HSCR^11^ ^12^. Altogether, the specific mechanism of HSCR remains elusive.

Remodeling of ENCs can occur postnatally. ENC loss and injury have been observed in patients with necrotizing enterocolitis^13^ and inflammatory bowel disease^14^ ^15^. Postnatal herpes simplex virus 1 infection promotes neutrophil-mediated destruction of the ENCs and HSCR-like megacolon in mice^16^. Given that the intestinal microenvironment changes dramatically after birth^4^, it is essential to determine whether abnormal intestinal factors contribute to disease aggregation postnatally. Currently, the cellular heterogeneity of human ENCs have been studied by spatial and single-cell transcriptomic sequencing^17–20^, but definitive characterization of ENCs in HSCR is lacking. Here, we present the cellular landscape of the intestinal ganglia and hypertrophic nerve trunks (HNTs) in patients with HSCR at single cell resolution and spatially. We show that intestinal ischemia is an important pathogenic mechanism (and potential therapeutic target) for infants with HSCR.

## MATERIALS AND METHODS

### Participants

The study procedures were approved by the Medical Ethics Committee of Guangzhou Women and Children’s Medical center (ID: 2020-42601), and complied with the international ethical guidelines for research involving human subjects as stated in the Helsinki declaration. The legal guardians of all patients assigned written informed consents. 36 age-matched healthy donors (H-CT), 28 anorectal malformations patients (CT), and sporadic HSCR patients (n=172) were recruited. Colonic tissues from 14 HSCR patients and 8 CT patients were used for constructing single-cell RNA transcriptome and spatial transcriptome. Whole exome sequencing was performed for 14 HSCR patients whose colonic biopsies were used for scRNA-seq. Bulk RNA-seq was applied to colonic tissues collected from a separated sub-cohort of subjects containing 60 HSCR patients and 13 CT patients. Participant details including developmental stage and sex were available at **online supplemental table 1. The random ID (Masked_ID) cannot reveal the identity of the study subjects and was not known to anyone outside the research group.**

### Single-cell RNA sequencing

Fresh tissues were processed with Chromium Next GEM Single Cell 5’ Kit v2 (10x Genomics, Cat# 1000263) following manufacturer’s instructions. The libraries were sequenced with Illumina Novaseq 6000. Cell Ranger (v.6.1.1) was used to process the sequencing results. The outputs of Cell Ranger were processed by scanpy package^21^ ^22^ (v.1.7.1). The cell differentiation trajectory was predicted by Monocle 3^23^ ^24^ (v.1.0.0). The cell-cell interactions were predicted by CellChat^25^ (v.1.1.0).

### Spatial transcriptome sequencing

Frozen tissues were processed with Visium Spatial Tissue Optimization Slide & Reagents Kit (10x Genomics, Cat# 1000193) and Visium Spatial Gene Expression Slide & Reagents Kit (10x Genomics, Cat# 1000187) following the manufacturer’s instructions. The cDNA libraries were sequenced with Illumina Novaseq 6000. Spaceranger (v.1.3.1) was used to process the sequencing results. The outputs were processed with the Seurat package^26^ (v.4.0.1).

### Bulk RNA sequencing

RNA was extracted from frozen tissues using FreeZol Reagent kit (Vazyme, Cat# R711-01). NEBNext Ultra II RNA Library Prep Kit for Illumina (New England Biolabs Inc, Cat# E7775) was used to construct cDNA libraries which were sequenced with Illumina Novaseq 6000.The raw reads were filtered by fastp^27^ (v.0.23.2) and aligned to human genomics using STAR^28^ (v.2.7.4a). CIBERSORTx^29^ were used to estimate the abundance of spatial transcriptome clusters in bulk RNA-seq data. The gene set score was calculated by Seurat package.

### Pathway enrichment

Metascape^30^ (https://metascape.org/) with default parameters were used for pathway enrichment. DEGs with adjusted *p* value lower than 0.05 and absolute value of log_2_(fold change) larger than 0.4 were used for pathway enrichment. Up-regulated and down-regulated DEGs were analyzed separately.

### Whole exome sequencing

DNA was extracted from blood using QIAamp DNA Mini Kit (Qiagen, Cat# 51304). Libraries were built using NEBNext® Ultra™ II DNA Library Prep Kit for Illumina® (NEB, Cat# E7645L) and SureSelectXT Human All Exon V6 (Agilent Technologies, Cat# 54898). NovaSeq 6000 was used for sequencing. The sequencing results were filtered by fastp^27^ (v.0.23.2) and were processed according to “GATK Best Practices” pipeline, in which bwa ^31^ (v.0.7.17), samtools^32^ (v.1.13), picard^33^ (v.2.21.4), and GATK^34^ (v.4.2.0.0) were used. ANNOVAR^35^ (v.2020-06-08) and VEP^36^ (v.104) were used to annotate the variants.

### Mice

C57BL/6 mice were purchased from Charles River (Beijing, China). *Sox10*^WT/MUT^ C3H mice were purchased from the Jackson lab (Cat# 000290). All mice were maintained in specific-pathogen-free (SPF) facility at the Guangzhou Medical University. The experimental protocols of this study were approved by the Ethics Committee of Guangzhou Medical University (ID: 2021-059B00). All experiments were performed in accordance with procedures outlined in the “Guide for Care and Use of Laboratory Animals” (National Resource Council, National Academy Press, Washington D.C).

### Establishment of the neonatal ischemic enterocolitis (NEI) mouse model

Neonatal ischemic enterocolitis was induced in 5-day-old mice of either gender, which were randomly divided into control and experimental groups, by gavage feeding newborn mice with enteric bacteria obtained from SPF wild type C57BL/6 mice or *Sox10*^WT/MUT^ C3H mice once per day for the first and second day of the experiment. Additionally, the mice were subjected to hypoxia (5% O_2_-95% N_2_) for 2 min in a hypoxia chamber (STEMCELL) twice daily and followed by 4 °C treatment for 10 min from the third to fifth day. Mice were then sacrificed for histopathological analysis. Colon tissues were collected for further analyses.

### Cell culture

For MEF isolation, mouse embryos (12.5-14 d) dissected out of head and red organs were washed with PBS, finely minced and treated with 1 ml of 0.05% trypsin EDTA (Gibco, Invitrogen) for 15 min at 37 °C. Isolated MEF cells were cultured in high-glucose (4.5 g/L) Dulbecco’s modified Eagle’s medium (DMEM, Gibco, San Diego, CA) supplemented with 10% fetal bovine serum (FBS, Hyclone, Logan, UT), 100 U/mL penicillin, 100 μg/mL streptomycin (Invitrogen Life Technologies), 15 mM 4-(2-hydroxyethyl)-1-piperazineethane sulfonic acid (HEPES) (pH 7.4, Invitrogen Life Technologies, San Diego, CA) complete medium.

SH-SY5Y cells (ATCC) were cultured in a 1:1 mixture of ATCC-formulated Eagle’s Minimum Essential Medium (ATCC) and F12 medium (Gibco, San Diego, CA) supplemented with 10% FBS, 100 U/mL penicillin, 100 μg/mL streptomycin (Invitrogen Life Technologies), 15 mM HEPES (pH 7.4, Invitrogen Life Technologies, San Diego, CA).

All cells were incubated in humidified atmosphere of 95% O_2_ and 5% CO_2_ at 37 °C incubator (Thermo Fisher Scientific). As for ischemia stress, cells were cultured in DMEM without glucose (Gibco, San Diego, CA) at 37 °C incubation with 5% CO_2_, 1% O_2_ and balanced N_2_.

### Quantification and statistical analysis

Statistical analysis of DEGs identification was performed by two-sided Mann-Whitney U test and bonferroni correction using all genes in the dataset. Two-side Mann-Whitney U test was also employed for the statistical analysis of gene set score. Statistical analysis of pathway enrichment was performed by hypergeometric test and Benjamini-Hochberg *P*-value correction algorithm. Statistical analysis and graphs of other data were generated using GraphPad Prism v9. Pairwise comparisons were analyzed using two-tailed unpaired Student’s *t*-test and multiple comparisons were analyzed with one-way analysis of variance (ANOVA). For all graphs and heatmaps, *P < 0.05, **P < 0.01, ***P < 0.001, and ****P < 0.0001. 95% confidence interval for all statistical analysis.

Detailed description of analysis pipeline, antibodies, and experimental protocols were available at **online supplemental methods**.

## RESULTS

### Cohort characteristics, single-cell and spatial transcriptomic profiling of the colonic microenvironment for infants with HSCR

In this study, we assembled 3 cohorts: age-matched healthy controls (H-CT, n=36), control subjects with anorectal malformations but normal ENC development (CT, n=28), and sporadic HSCR patients (n=172). Clinical manifestations and major laboratory findings for the participants are presented in **figure 1A** (**online supplemental table 1**). HSCR patients were mainly males (80.8%), the majority of whom had abdominal distention (89.5%), vomiting (66.3%), and delayed meconium passage (74.4%). Immunological workup revealed significantly increased circulating platelet and lymphocyte counts in HSCR compared with H-CT and CT subjects.

**Figure 1.**
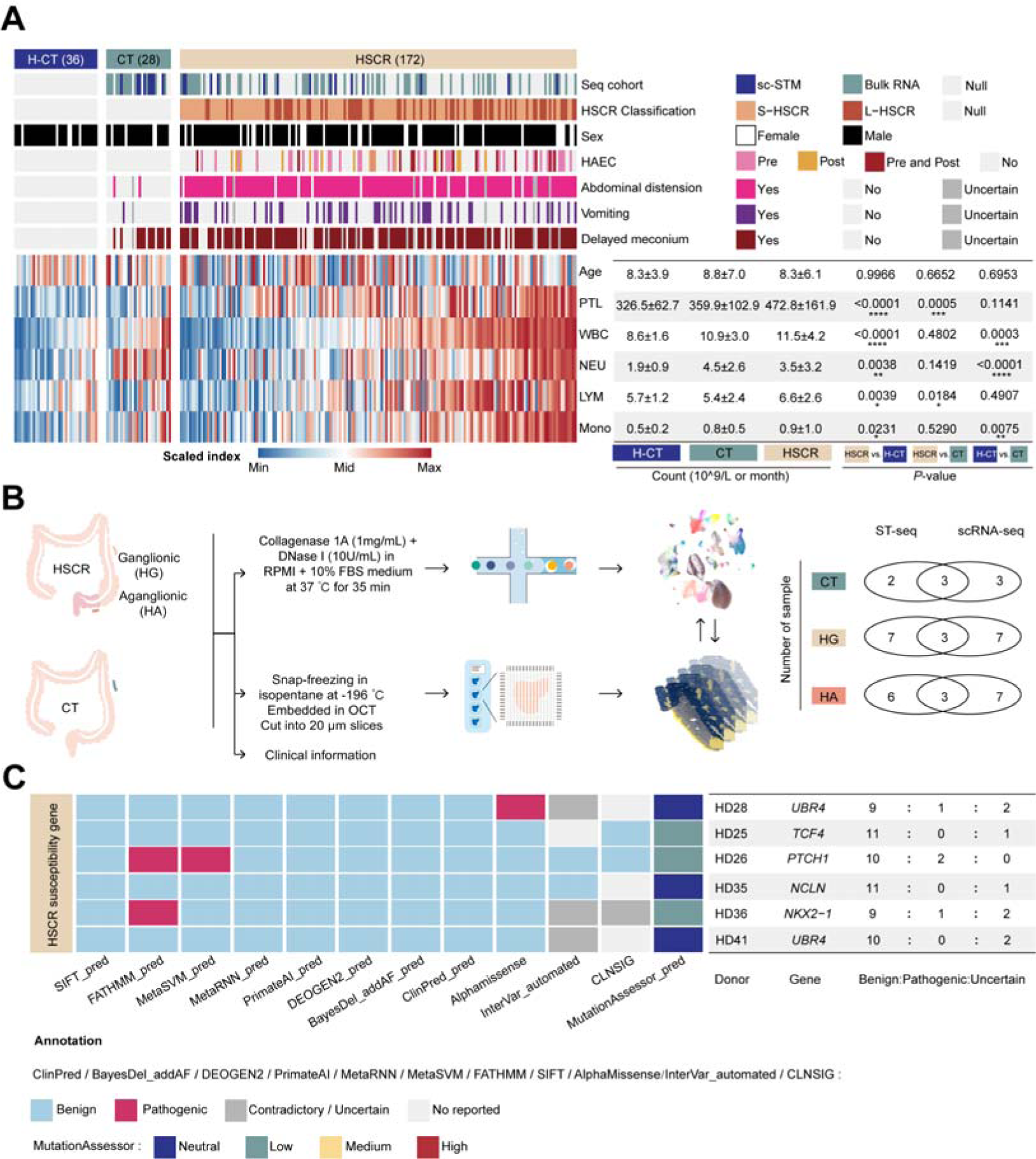
Cohort characterization. **(A)** 36 age-matched healthy donors (H-CT), 28 anorectal malformations patients (CT) and 172 sporadic HSCR patients were recruited in this study. Heatmap displaying clinical information recorded on the electric medical records. Among these donors, scRNA-seq and ST-seq were applied to colonic tissues from eight CT and 14 HSCR subjects, bulk RNA-seq was applied to colonic tissues from a different set of 13 CT and 59 HSCR donors (HG=53, HA=54; 48 HG-HA pairs). The mean, standard deviation (SD), and *P* value were shown in the table. Two-tailed *t*-test. **** *P* < 0.0001, \*\*\**P* < 0.001, \*\**P* < 0.01, \**P* < 0.05. S-HSCR, short segment HSCR. L-HSCR, long segment HSCR. Delayed meconium, inability to pass meconium within the first 48 hours of life. PTL, blood platelet counts. WBC, blood white blood cell counts. NEU, blood neutrophil counts. LYM, blood lymphocyte count. Mono, blood monocyte counts. Colors were used to show the normalized order of specific parameter values. **(B)** Workflow for sample collection and data analysis. Sample numbers are indicated. **(C)** Grid plot showing rare genetic variants of known susceptibility genes detected in 14 HSCR patients applied with scRNA-seq and ST-seq. Each row represents a variant. All variants are heterozygous single nucleotide variants with a total allele frequency (according to dbSNP) less than 1%. 12 databases were used to annotate these variants.

We generated spatial and single-cell transcriptomes for 8 CT and 14 HSCR patients **(figure 1B)**. We also performed whole exome sequencing for the 14 HSCR patients. For independent validation, we generated bulk RNA sequencing using colonic biopsies from a separate sub-cohort of CT (n=13) and HSCR patients (aganglionic HSCR, HA, n=54; ganglionic HSCR, HG, n=53.) (**figure 1A**). We analyzed the rare genetic variants from the 14 HSCR patients against the list of candidate HSCR risk genes (**online supplemental table 2**), and no known pathogenic variant was identified (**figure 1C and online supplemental table 3**). This was consistent with the sporadic nature of our HSCR cohort.

For spatial transcriptomes, we obtained 50,234 spots (ST-spots) and partitioned them into 13 clusters (ST-clusters) using a semi-supervised analysis (**see Method**). ST-clusters exhibited transcriptional signatures that are consistent with their spatial distributions (**figure 2A, 2B, and online supplemental figure 1A-1D**). For scRNA-seq, we partitioned 193,800 high-quality cells into 61 unique populations (**online supplemental figure 1E and 1F**) according to unsupervised clustering and known marker genes (**online supplemental figure 2 and online supplemental table 4**). ENCs were not captured in our scRNA-seq dataset due to technical limitations. Therefore, we included a published single-nucleus transcriptome dataset for ENCs in our analysis^37^.

**Figure 2.**
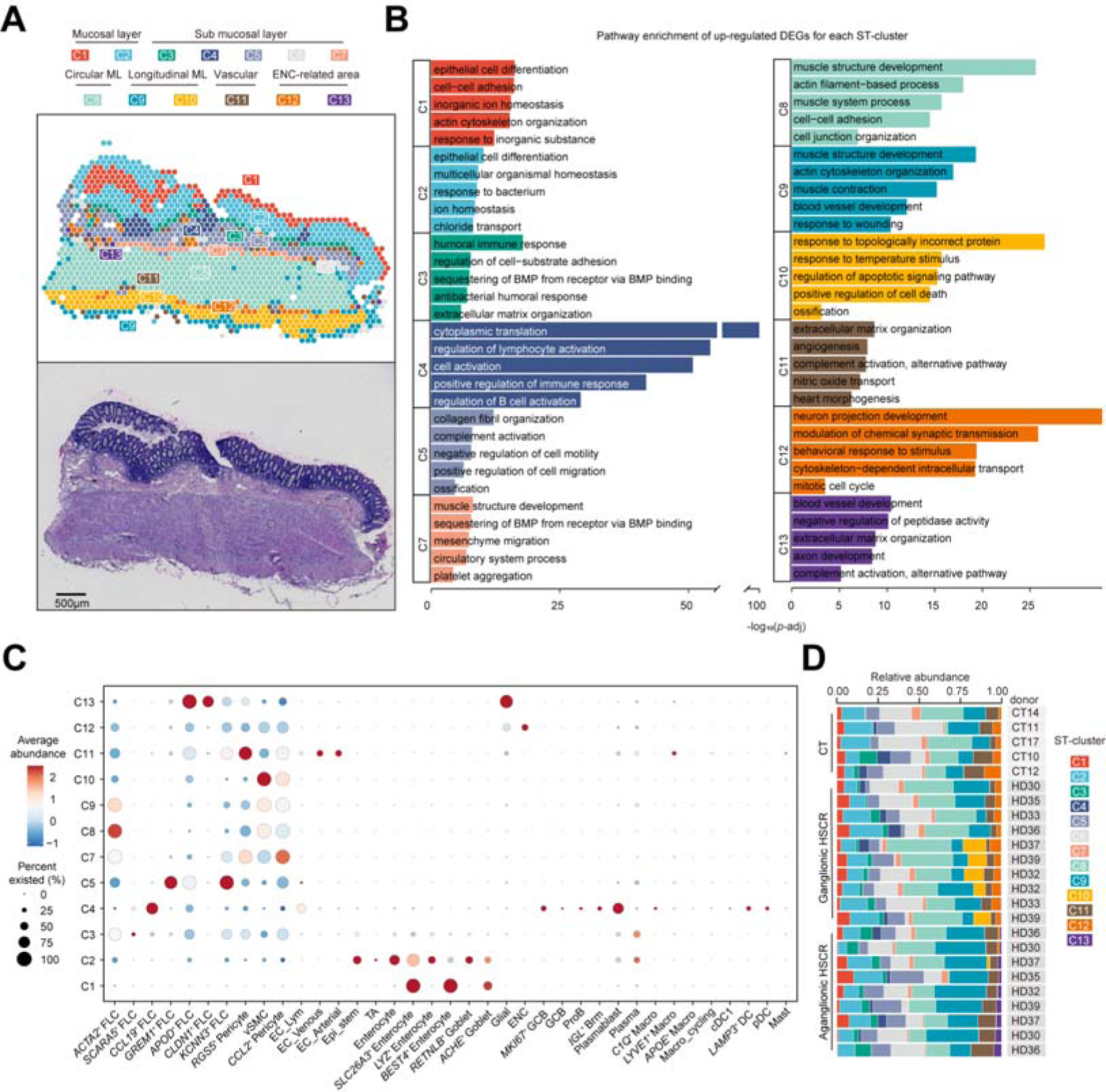
Single-cell and spatial transcriptome profiling of HSCR and control subjects. **(A)** Distribution of 13 spatial transcriptome (ST) clusters (C1-C13) in a representative ST slide. **(B)** Bar plot displaying pathways enriched in up-regulated DEGs of each ST cluster. **(C)** Heatmap showing abundances of scRNA-seq subsets on ST-clusters. Color represents center-normalized relative abundance. The size of dots represents the fraction of ST-spots containing corresponding cells. **(D)** Bar plot showing the relative abundance of ST clusters for control and HSCR subjects.

For combined ST-seq and scRNA-seq analysis, we utilized an anchor-based workflow to link scRNA-seq-defined cell populations with ST-spots (**see Method**). This allowed spatial alignment of 37 scRNA-seq populations on the colonic biopsies (**figure 2C**). For example, *BEST4*^+^ enterocytes were located at the top of the mucosa (C1), undifferentiated epithelial cells (TA and Epi_stem) at the base of the mucosa (C2), pericytes and endothelial cells near blood vessels (C11), and immune cells at the mucosal-associated lymph follicles (C4). Other scRNA-seq-defined cell populations have diffused distribution patterns and were unable to be defined spatially.

### Identification of differentially enriched ST-clusters and cell types in HSCR

By comparing HSCR with control subjects, we identified 3 unique ST-clusters (**figure 2D**). ST-cluster C12, which contained ENC-related signatures, were absent in the aganglionic HSCR (HA) colons. ST-cluster C13, encompassing glial cells, *CLDN1*^+^ fibroblast-like cells (FLCs) and *APOD*^+^ FLCs, was only identified in HA colons. ST-cluster C10, a muscular-associated cluster enriched with cellular stress-related pathways, was present in 5 HG and 1 HA colons but was absent in CT colons. (**figure 2A-2D)**. At the single cell level, our data showed that many immune cells, including Th17, macrophages and plasma cells, were significantly increased in the colons of patients with HSCR compared with control subjects **(online supplemental figure 1F)**. This potentially contributes to enterocolitis risk in patients with HSCR^38^.

### Analysis of muscular associated spatial cluster C10 reveals prevalence of ischemic stress in HSCR

We first analysed C10, a muscular ST-cluster identified only in infants with HSCR (**figure 2D**). Bulk RNA-seq data deconvolution confirmed that C10 was significantly increased in HG and HA colons compared with the control subjects (**figure 3A**). To examine whether C10 may represent a transitional or intermediate disease state, we compared C10 with the rest of the muscular regions (C8, C9) (**figure 3B, 3C, and online supplemental figure 3A**). The upregulated genes in C10 included myocardial infarction-related transcription factors (such as *FOSL1* and *THBS1*)^39^, genes that mediate misfolded protein responses, chemotaxis, and response to zinc ion. The downregulated genes encode collagens (*COL1A1, COL1A2, COL4A1*) and regulate muscular contraction (*ACTG2*, *CALD1*, *MYH11*). These changes resemble cellular responses to ischemic stress^39^. Indeed, analysis of the ischemic response gene set showed that C10 had the highest ischemia score compared with the rest of the muscular regions (**figure 3D**). At the cellular level, C10 contained increased numbers of vascular smooth muscle cells (vSMC) and ischemia-related pericytes (*CCL2*^+^ Pericyte) but had decreased myofibroblast (*ACTA2*^+^ FLCs) cells (**figure 3D and online supplemental figure 3B-3E**).

**Figure 3.**
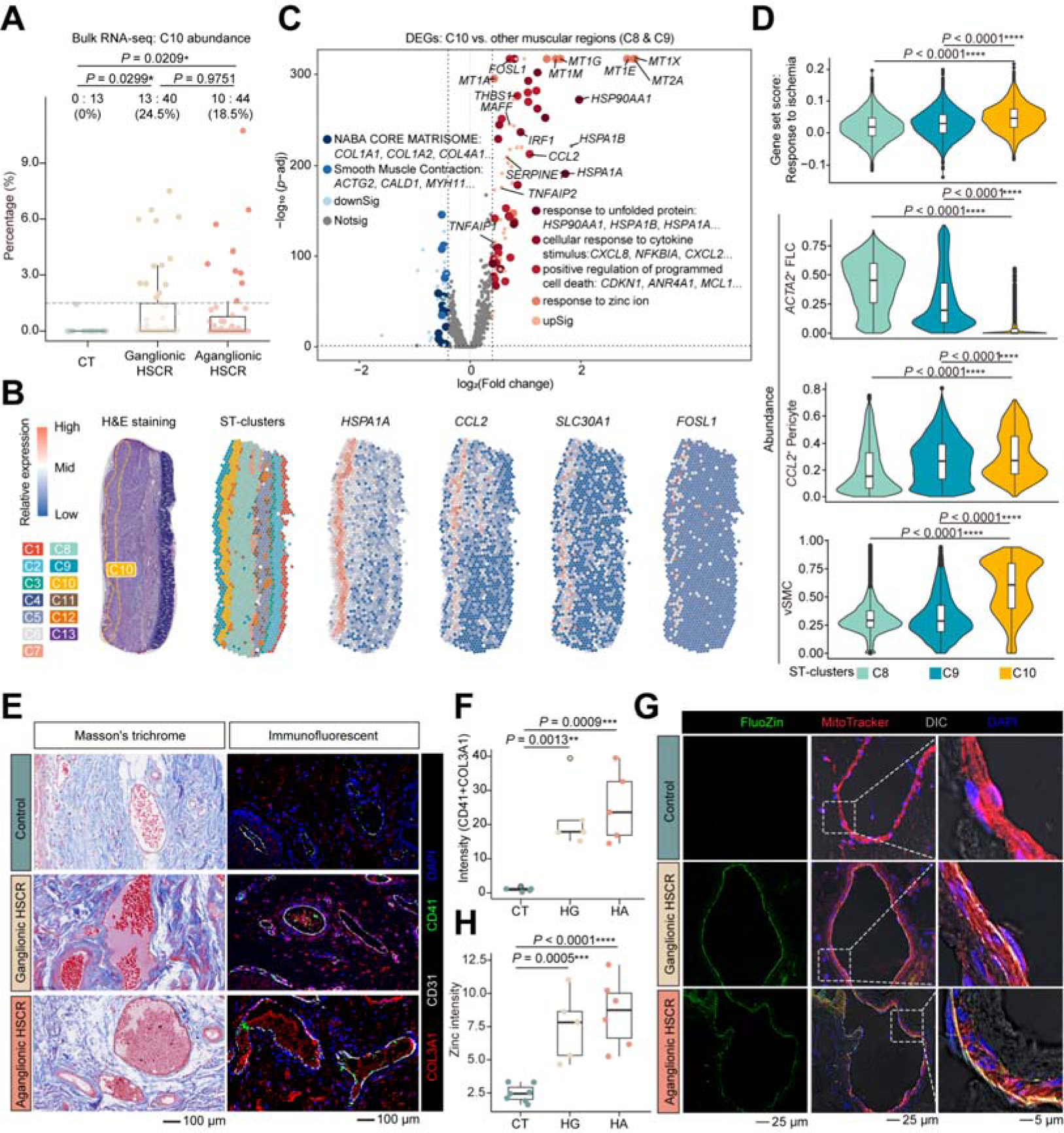
Ischemic stress in HSCR colons. **(A)** Box plot showing the abundance of C10 imputed by CIBERSORT in bulk RNA-seq data. The percentage of HG and HA samples with a C10 abundance higher than the confidence limit (1.5%) was marked. **(B)** The expression of selected genes on a representative slide of HG colon. **(C)** Volcano plot showing DEGs in the comparison between C10 and the rest of the muscular ST-clusters (C8 and C9). DEGs of selected pathways were indicated as colored bold spots. **(D)** Violin-box plot showing gene set score (response to ischemia, GO: 0002931) and the abundance of single-cell populations in the muscular ST-clusters. (**E**-**F**) Masson’s trichrome and immunofluorescent imaging (**E**), and box plot (**F**) showing platelet and collagen aggregates in the colonic blood vessels (five donors for each group). (**G-H**) Zinc ion (Fluozin) and mitochondria (MitoTracker) staining (**G**) and box plot (**H**) showing the concentration of zinc ion in the colonic vasculatures (seven CT, five HG, and six HA subjects). Each datapoint in the same group represents a donor (**A**, **F**, **H**). For boxplots, line within box represents the median; the box limit represents the SD, the whiskers represent the 1.5 × outliers (**A**, **D**, **F**, **H**). Statistical significance was determined using two-sided Mann-Whitney U test (**A, D**) or two-tailed *t*-test (**F**, **H**). \*\*\*\**P* < 0.0001, \*\*\**P* < 0.001, \*\**P* < 0.01, \**P* < 0.05.

Activated platelets are known to infiltrate ischemic tissues and form microthrombi composed of platelets and collagens^40^. Zinc is released from the platelets and further exacerbates thrombosis^41^. We found platelet-collagen aggregation in the vasculature of the HG and HA colons but not in the control subjects (**figure 3E and 3F**). Zinc-dye staining (FluoZin) showed that zinc ion was increased significantly in the vascular walls in HSCR compared with the control colons (**figure 3G and 3H**). Consistently, the score of genes that corresponded to cellular responses to zinc ion were significantly up-regulated in vascular endothelial cells in HSCR compared with control colons (**online supplemental figure 3F**).

### Identification of HNT-associated *CLDN1*^+^ FLCs in HSCR

HNTs (C13) consisting of glial cells and ENC fibers, is a feature of HSCR^11^. To reveal mechanisms underlying HNT formation, we compared HNTs (C13) with ganglia (C12). Genes that are expressed by glial cells (*GPM6B*, *MPZ* and *GFRA3*) and fibroblast-like cells (*COL3A1*, *COL1A1*) (**online supplemental figure 2**), and genes that regulate blood vessel development (*APOD, ANGPTL7, NOTCH3*), were significantly enriched in HNTs compared with ganglia (**figure 4A, 4B, and online supplemental figure 4A**). In contrast, genes that are expressed by ENCs (*VIP*, *MAP2*) (**online supplemental figure 2**), genes that regulate neuron projection development (*GAP43, NPY, RET*), and genes that regulate ATP metabolism (*ATP1B1*, *ATP5F1A, ATP5F1E*) were significantly down-regulated in HNTs compared with ganglia (**figure 4B and online supplemental figure 4A**).

**Figure 4.**
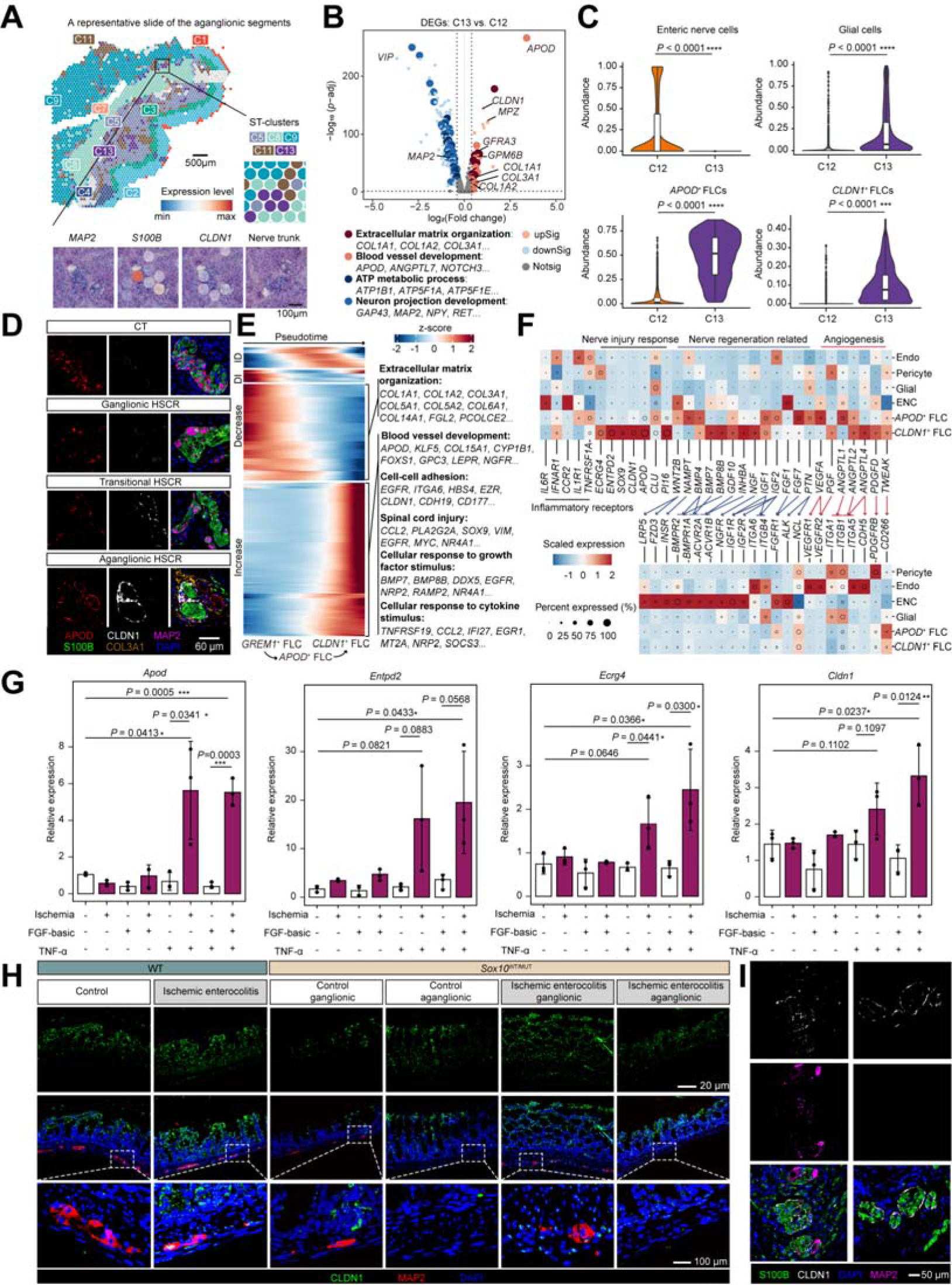
Ischemic stress, FGF and TNF-α induce peri-neural and peri-HNT *CLDN1*^+^ FLCs. **(A)** A spatial map displaying ST clusters (upper panel) and expression of selected genes in HA colons (lower panel). Normalized expression levels were presented by color. **(B)** Volcano plot displaying the DEGs between ganglia (C12) and HNTs (C13). DEGs of selected pathways were marked with bold spots. **(C)** Violin-box plot showing the abundance of scRNA-seq subsets in ganglia and HNTs, respectively. Two-sided Mann–Whitney U test. Data are median (line within the box) with a SD (box limit) and 1.5 × outliers (whiskers). **(D)** IF imaging (six donors for each group) showing compositions of ganglia and HNTs in control and HSCR colons. **(E)** Heatmap displaying transcriptional changes during the differentiation of indicated FLCs. **(F)** Heatmap showing the expression of selected ligands and receptors in scRNA-seq defined subsets. Color represents center-normalized gene expression level. The size of dots represents the fraction of cells expressing corresponding genes. Ligand-receptor pairs were linked by arrows. **(G)** Bar plot showing the relative expression of *CLDN1*^+^FLC marker genes in MEFs that were cultured under indicated conditions. Three biological replicates are expressed as mean ± SD. Two-tailed *t*-test. Each data point represents an independent assay. \*\*\**P* < 0.001, \*\**P* < 0.01, \**P* < 0.05. **(H)** IF imaging (five animals for each group) showing CLDN1 and MAP2 staining in wild type (WT) and *Sox10*^WT/MUT^ mice under indicated conditions. **(I)** IF imaging (representative results from three donors) displaying HNT-like structures in the colons of patients with Crohn’s disease.

At the cellular level, we confirmed that HNTs contained significantly increased numbers of glial cells, *CLDN1*^+^ FLCs, and *APOD*^+^ FLCs than ganglia while ENCs were absent in HNTs (**figure 4C**). Notably, *CLDN1*^+^ FLCs were detected almost exclusively in HA colons (**online supplemental figure 1F**). By Immunofluorescence (IF) imaging, we found that *CLDN1*^+^ FLCs wrapped the HNTs and that scattered *CLDN1*^+^ FLCs occurred around some ganglia in the transitional zones between the HG and HA colons (**figure 4D)**. Trajectory analysis suggested that *CLDN1*^+^ FLCs were differentiated from *APOD*^+^ FLCs, and that trophocytes (*GREM1*^+^ FLCs)^42^ were the common progenitors for both FLCs (**figure 4E and online supplemental figure 4B-4C)**. Following differentiation, *CLDN1*^+^ FLCs lose the capacity for extracellular matrix organization, but acquire transcriptional programs that promote blood vessel development, cell-cell adhesion, growth factor responses, cytokine responses and neuron injury responses (**figure 4E and online supplemental figure 4D-4E)**. For example, apolipoprotein D (encoded by *APOD)* is an injury-associated peripheral neuron regeneration factor^43^ and Claudin 1 (encoded by *CLDN1)* is an integral membrane protein induced at the leaky blood-brain barrier of patients with chronic stroke^44^.

Cell-cell interaction analysis suggested that *APOD*^+^ FLCs and *CLDN1*^+^ FLCs may promote angiogenesis by expressing *VEGFA*, *PGF*, and *ANGPTL1*/*2*/*4* **(figure 4F and online supplemental figure 4F)**. IF imaging confirmed that endothelial cells around the HNTs in HA colons are increased compared with those around the ganglia in HG and CT colons (**online supplemental figure 4G)**. *APOD*^+^ FLCs and *CLDN1*^+^ FLCs may also regulate the fate of ENCs and glial cells by secreting neuron regeneration factors, including WNT2B^45^ ^46^, bone morphogenetic proteins (BMPs)^47^, NAMPT^48^, NGF^49^, insulin-like growth factors (IGFs)^50^, GDF10^51^ and FGF7^52^ (**figure 4F)**. Our analysis also suggested that ENCs may play a role in the differentiation of *APOD*^+^ FLCs and *CLDN1*^+^ FLCs via the FGF1-FGFR1 signalling (**figure 4F)**. In addition, we found that TNFR1 (encoded by *TNFRSF1A*), the receptor for the inflammatory cytokine TNF-α, was expressed by the *APOD*^+^ FLCs and *CLDN1*^+^ FLCs **(figure 4F)**.

### Ischemia, inflammation and ENC promote *CLDN1*^+^ FLCs development in HSCR

From the above analysis, we hypothesized that ischemic stress, inflammation and neuron-fibroblast interactions may promote the differentiation of *CLDN1*^+^ FLCs. Therefore, we established an *in vitro* assay using mouse embryonic fibroblasts (MEFs). The expression of mouse homologues of *CLDN1*^+^ FLCs marker genes (*Cldn1*, *Apod*, *Ecrg4*, *Entpd2*) were significantly up-regulated when MEFs were cultured under ischemic conditions (12 hr glucose and oxygen deprivation) in the presence of TNF-α and FGFR1 ligand (FGF-basic) (**figure 4G**).

To validate these findings *in vivo*, we established a HSCR model of neonatal ischemic enterocolitis (NIE) using C57BL/6 mice (**online supplemental figure 5A-5F**). Similar to HSCR patients, we found that NIE mice had a C10-like muscular layer (TNF-α^+^ and CCL2^+^) (**online supplemental figure 5G)**, HNTs (**online supplemental figure 5H)**, and peri-ganglia CLDN1 expression (**figure 4H**). To further confirm the requirement of neuron-derived signals for *CLDN1*^+^ FLCs differentiation, we studied *Sox10*^WT/MUT^ C3H mice, which are known to have defective ENCC migration to the distal colon^53^ (**online supplemental figure 6A-6G)**. CLDN1 was detected in the peri-ganglionic regions of the *Sox10*^WT/MUT^ NIE mice but not in the *Sox10*^WT/MUT^ control mice (**figure 4H**). Consistent with the requirement of neuron-derived signals, peri-ganglia CLDN1 and HNTs were absent in the aganglionic colons in the neonatal *Sox10*^WT/MUT^ mice (**figure 4H and online supplemental figure 6C**).

Altogether, our results suggest that ischemic stress promotes *CLDN1*^+^FLC differentiation and HNT formation independently of ENCC migration defects.

### Ischemia and inflammation promote *CLDN1*^+^FLCs and HNT formation in patients with Crohn’s disease

As ischemia is found in many inflammatory diseases of the intestine, we foreshadowed that HNT-related findings for HSCR might also occur these other diseases. Patients with Crohn’s disease (CD) have chronic and relapsing intestinal inflammation and ischemia^54–57^. To determine whether the mechanism we showed above may also regulate ENCs in CD, we examined surgical biopsies. We found that scattered CLDN1 nearby ganglia and CLDN1-wrapped HNTs are also extensively present in CD colons (**figure 4I**).

### Ischemia induces ENC death in HSCR

To determine whether ischemia also contributes to the loss of ENCs in HSCR, we examined the ganglia (C12). A subgroup of inflammatory muscular ganglia (iMus_ganglia) was identified in 3 of 7 HSCR patients **(figure 5A-5D)**. Compared with the noninflammatory muscular ganglia (nMus_ganglia), the iMusc_ganglia upregulated the expression of inflammatory cytokines (*TNF*, *CCL2*), displayed increased response to zinc ions, and downregulated the activity of synaptic transmission **(figure 5E)**. Gene set score analysis showed that neuron apoptosis pathway was significantly increased in the iMusc_ganglia compared with the nMusc_ganglia **(figure 5F)**. Analysis of bulk RNA-seq data confirmed the increased apoptosis score in HSCR compared with control subjects **(figure 5G)**. By fluorescent imaging, we confirmed that zinc ion and zinc ion response proteins (SLC30A1 and MT1A) were significantly increased in the ENCs in HG compared with the control colons **(figure 5H-5J)**. Furthermore, TNF-α and cleaved CASP3 (the apoptosis executor protein) were significantly increased in ENCs of the HG colon compared with the control colon **(figure 5K and 5L)**. Similarly, in the NIE mice, we also observed significantly increased CASP3 cleavage, mitochondrial cristae breakage, mitochondrial swelling, and nuclear deformation in the ENCs compared with the control ENCs (**online supplemental figure 5I and 5J**).

**Figure 5.**
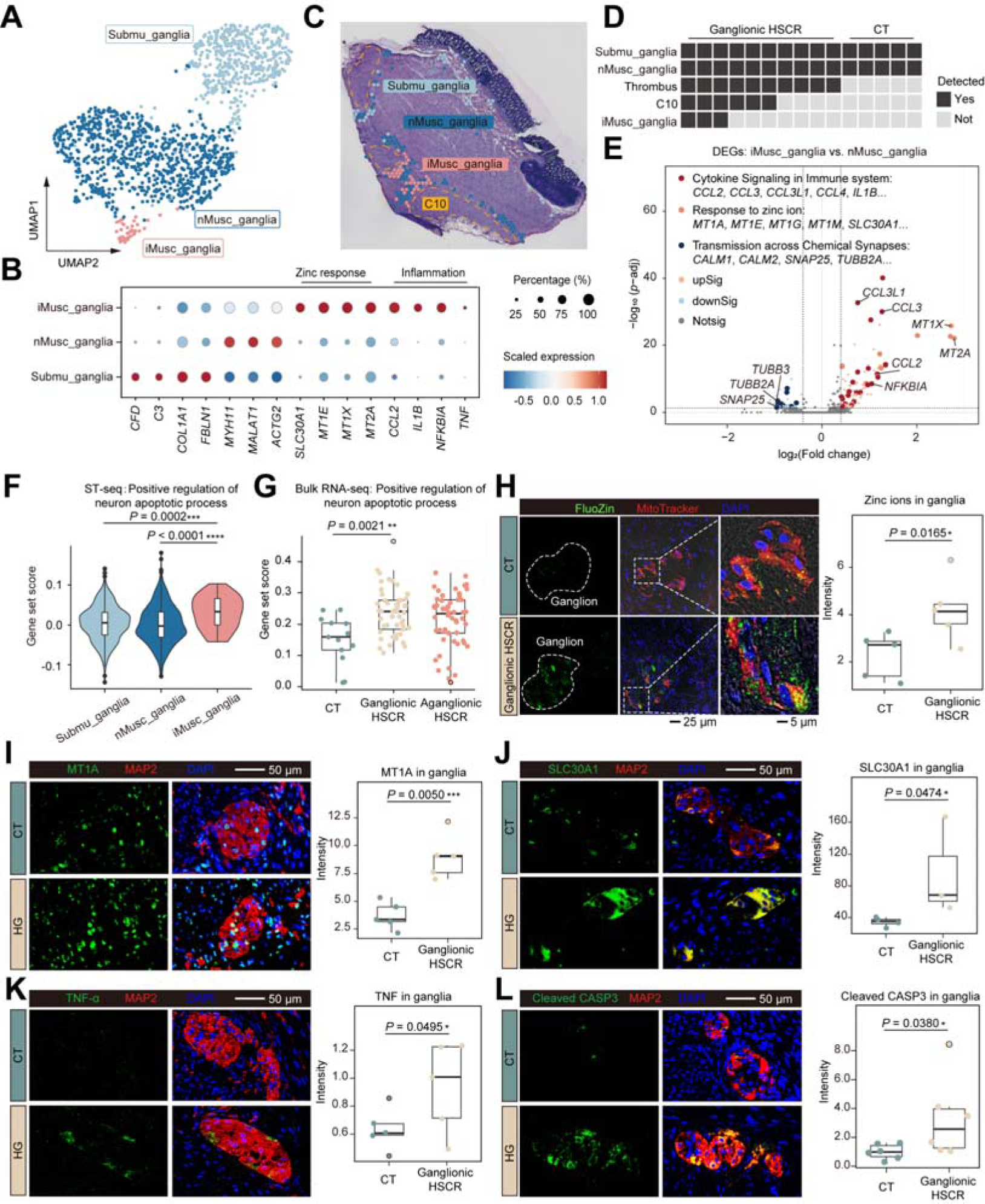
Neuroinflammation and apoptosis in HSCR colons. **(A)** UMAP plot showing the C12 (ganglia) sub-clusters. Submu_ganglia, ST-spots that mainly cover ganglia in submucosa region. nMusc_ganglia, ST-spots that mainly cover non-inflamed ganglia in muscular region. iMusc_ganglia, ST-spots that mainly cover inflammatory ganglia in muscular region. **(B)** Heatmap displaying the expression of marker genes for ganglia sub-clusters. The colors represent the center-normalized relative expression levels of genes. The size of dots represents the fraction of ST-spots expressing corresponding genes. **(C)** An H&E slide from an HG colon showing the location of the ganglia sub-clusters. **(D)** Grid plot showing co-appearance of C10 and ganglia sub-clusters in HSCR. Each square represents a ST-seq sample. **(E)** Volcano plot showing DEGs of the comparison between nMusc_ganglia and iMusc_ganglia. DEGs of the selected pathways were marked with colored thick spots. **(F)** Violin-box plot showing the gene set score (GO: 0043525, positive regulation of neuron apoptotic process) in ganglia sub-clusters. **(G)** Box plot showing the gene set score (GO: 0043525, positive regulation of neuron apoptotic process) in bulk RNA-seq. (**H-L**) Fluorescence imaging and boxplot displaying zinc ion, MT1A, SLC30A1, TNF-α, and cleaved CASP3 in the ganglia from control and HSCR subjects. Data are median (line within the box) with a SD (box limit) and 1.5 × outliers (whiskers) (**F-L**). Statistical analysis was performed using two-sided Mann-Whitney U test (**F-G**) or one-tailed *t*-test (**I-L**). \*\*\*\**P* < 0.0001, \*\*\**P* < 0.001, \*\**P* < 0.01, \**P* < 0.05. Each datapoint within the same group represents a donor (**G-L**). Representative fluorescence images were generated of slides of the same number of donors shown in the boxplot (**H**, **I**, **K**: five HSCR and five CT; **J**: three HSCR and four CT; **L**: six HSCR and six CT).

To confirm that ischemia regulates ENC fate, we treated a human neuron cell line (SH-SY5Y) under ischemic conditions (6 hr glucose and oxygen deprivation). For SH-SY5Y cells, ischemia treatment significantly increased the expression of *TNF* and *CCL2*, promoted cellular zinc accumulation, mitochondria depolarization, and apoptosis (**figure 6A-6C**). Transmission electron microscope (TEM) revealed that ischemia-induced neuron apoptosis was associated with cytoplasmic shrinkage, membrane blebbing, mitochondrial damage and apoptotic body formation in SH-SY5Y cells **(figure 6D)**. Previously, ischemia has been shown to induce zinc ion accumulation and mitochondria damage^58^ ^59^. We confirmed that zinc ion chelation (TPEN), CASP3/8 inhibition, and ddC-mediated mitochondria DNA depletion^60^ significantly prevented SH-SY5Y from ischemia-induced apoptosis **(figure 6B and online supplemental figure 7A-7F)**.

**Figure 6.**
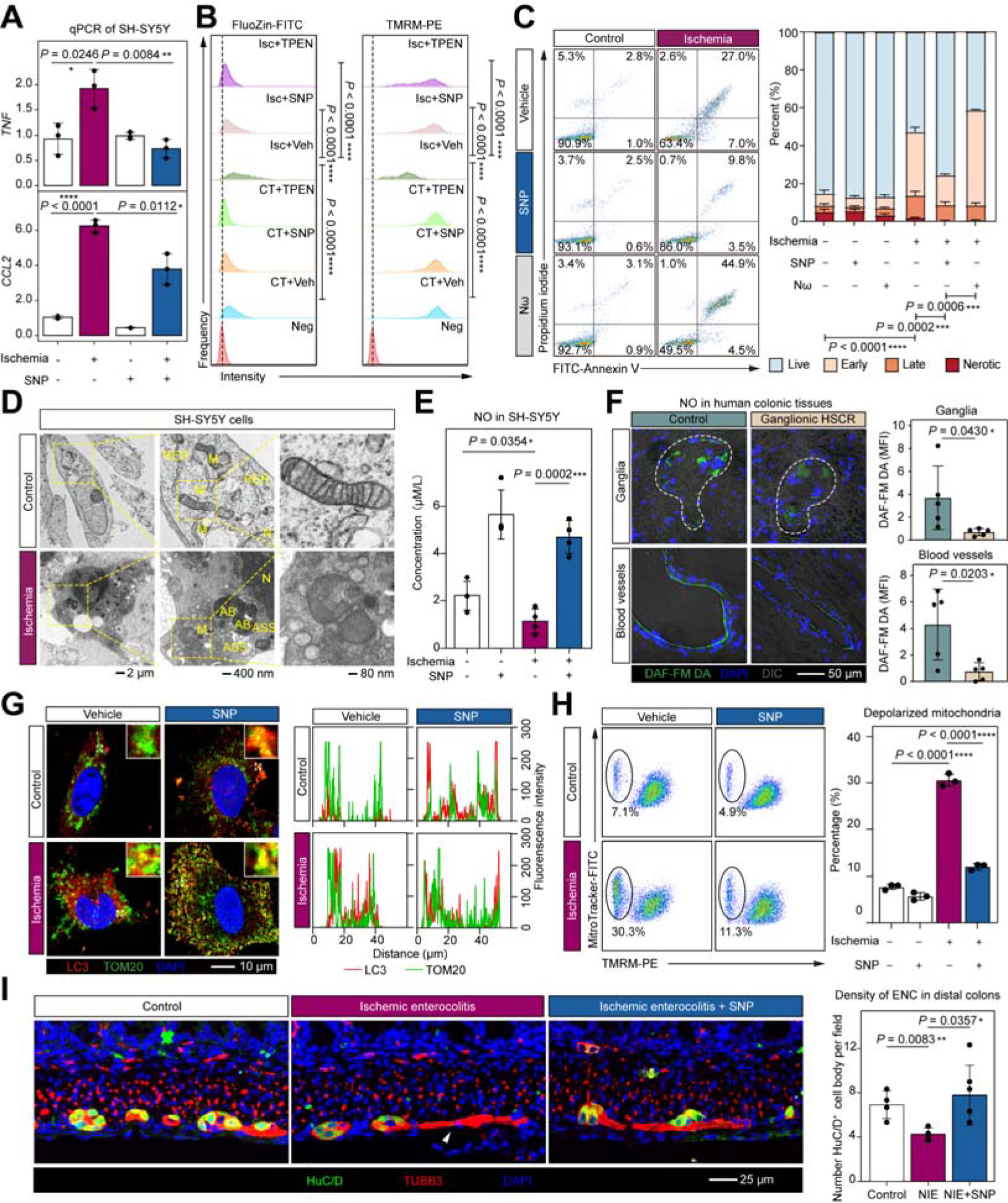
Nitric oxide prevents ischemia-induced enteric neuron cell death. **(A)** Bar plot showing the expression of *TNF* and *CCL2* in SH-SY5Y cells under the indicated conditions. SNP, sodium nitroprusside, a NO donor drug. **(B)** Frequency plot displaying the changes of intracellular free zinc ions (left panel) and mitochondrial membrane potential (right panel) in SH-SY5Y cells under the indicated conditions. Veh, vehicle (DMSO). TPEN, a zinc ion chelating agent. Neg, negative control. Isc, ischemia. CT, control condition. FlouZin, a zinc ion fluorescent dye. TMRM, a mitochondrial membrane potential indicator. **(C)** Scatter plot and bar plot displaying the types of cell death under indicated conditions detected by flow cytometry. Nω, an nNOS inhibitor. Early apoptotic (Annexin V^+^PI^-^), late apoptotic (Annexin V^+^PI^-^), necrotic (Annexin V^-^PI^+^). **(D)** TEM showing the structures of nuclei (N), mitochondria (M), apoptotic bodies (ABs), rough endoplasmic reticulum (RER), and autolysosome (ASSs) of SH-SY5Y cells under the indicated conditions. **(E)** Bar plot displaying the changes of NO levels in SH-SY5Y cell under indicated treatments. **(F)** Fluorescence imaging and bar plot showing the NO levels in the blood vessels and ganglia of HG and control colons. Slides are from five donors for each group. DAF-FM DA, a reagent that detect nitric oxide. MFI, mean fluorescence intensity. **(G)** IF imaging and frequency plot showing the mitophagy in SH-SY5Y cell under indicated treatments. TOM20 represents mitochondria. LC3B, a marker of autophagic activity. **(H)** Scatter plot and bar plot displaying the changes of mitochondrial membrane potential in SH-SY5Y cell under indicated treatment. **(I)** IF imaging and bar plot showing the density of ENC body in the distal colons of control (four animals), NIE (four animals), and NIE+SNP mouse (six animals). NIE+SNP, NIE mouse treated with SNP (5 mg/kg). Data are mean ± SD, each datapoint represents an assay (**A**, **E**, **H**), a donor (**F**) or an animal (**I**). Statistical analysis was performed using two-tailed *t*-test (**A-C**, **E**, **F**, **H**, **I**). \*\*\*\**P* < 0.0001, \*\*\**P* < 0.001, \*\**P* < 0.01, \**P* < 0.05. n= three (**A-D**, **F-H**) or four (**E**) independent experiments.

### Nitric oxide ameliorates ischemia-induced ENC death in NIE mice

Nitric oxide (NO) inhibits platelet activation and promotes vasodilation^61^. Recently, we showed that NO also promotes removal of damaged mitochondria by inducing mitophagy^62^. Synthesis of nitric oxide is limited under ischemic conditions due to oxygen deprivation. In SH-SY5Y cells, we confirmed that ischemia decreased NO levels **(figure 6E)**. In patients with HSCR, we also found that NO levels were significantly decreased in the ganglia and vascular walls of HG colons compared with the control subjects **(figure 6F)**.

To investigate whether NO may prevent cell death by promoting mitophagy, we treated SH-SY5Y cells with sodium nitroprusside (SNP), a clinically available NO donor drug. SNP significantly increased cellular NO concentrations and mitophagy (LC3 colocalization with mitochondria) **(figure 6E, 6G, and online supplemental figure 7G)**. SNP also decreased inflammation, zinc ion accumulation, mitochondrial depolarization, and cell death in SH-SY5Y cells during ischemia **(figure 6A-6C, 6H, and online supplemental figure 7G)**. The effects of SNP on ischemic SH-SY5Y cells were diminished under ddC treatment **(online supplemental figure 7D-7F)**, confirming the major contribution of mitochondria. In contrast, inhibiting neuronal NO synthase activity by nitroarginine (N_ω_) increased the apoptotic effects of ischemia **(figure 6C)**. It should be mentioned that directly activating cGMP, another downstream effector molecule of NO^61^ ^63^, only marginally alleviated SH-SY5Y cell death under ischemic conditions **(online supplemental figure 7H-7J)**. To evaluate the therapeutic effects of the NO donor drug *in vivo*, we treated the NIE mice with SNP. SNP promoted neonatal growth and prevented ENC loss in the NIE model (**figure 6I and online supplemental figure 5K-5M**).

## DISCUSSION

In this study, we generated single-cell and spatial transcriptomes for the ganglionic and aganglionic colons of infants with HSCR. Analysis of a muscular-associated C10 cluster that present exclusively in HSCR infants reveal extensive ischemia-related stress responses in HSCR. The intestinal vasculatures were clogged by aggregated platelets and collagen. Ischemia, inflammation and neuron-derived signals promote the differentiation of *CLDN1*^+^FLCs, which lie at the periphery of the HNTs and support neuron regeneration and angiogenesis. However, under persistent oxygen deprivation, NO becomes a limiting factor for the ENCs, which succumb to mitochondria damage, neuron inflammation and Caspase-mediated apoptosis. Finally, the pathophysiological significance of ischemia-related neuropathy in HSCR was confirmed by the amelioration of neuron death *in vitro* and in the NIE mouse model of human HSCR *in vivo* by SNP, a NO donor drug.

Defective ENCC migration has been suggested as the underlying cause of HSCR^5^ ^64^ ^65^. This is consistent with the fact that most HSCR patients in our cohort presented symptoms (delayed meconium passage) immediately after birth. However, glial cells derived from ENCCs^3^ are broadly present in most of the HA colons^11^ ^12^, which indicates success colonization of ENCCs in the patients. Interestingly, glial cell-containing HNTs are absent in HSCR patients whose entire colons are aganglionic^66^ ^67^. Consistent with the above clinical observations, glial cells and HNTs are absent in the aganglionic colons in *Sox10*^WT/MUT^ mice, which are known to have ENCC migration defects^53^. By analyzing colonic tissues of sporadic HSCR infants, we show for the first time that ischemia contributes to ENC loss in HSCR colons.

Using *Sox10*^WT/MUT^ NIE model, we demonstrate that only in the presence of neuron-derived signals, ischemia promotes the formation of CLDN1-wrapped HNTs. Given that intestinal obstruction leads to intestinal ischemia^68^, our results support a pathophysiological cascade of HSCR development, which may involve obstruction, ischemia, selective loss of ENCs in ganglionic colons, and elongation of aganglionic colons.

Our findings highlight the importance of preventing ischemia-related ENC loss in infants with HSCR. As a vasodilator and an inhibitor of platelet activation, NO has been used to treat ischemia for decades^61^. Moreover, our previous work had identified a novel function of NO, which removes damaged mitochondria by inducing mitophagy^62^. By culturing a neuron cell line under ischemic conditions, we observed significantly increased mitochondria depolarization, mitochondria damage and caspase-dependent neuron cell death. The therapeutic effects of NO donors as candidate treatments for HSCR are supported by SNP restoring neonatal growth and preventing ENC loss in the NIE model. Altogether, our study reveals that ischemia-induced postnatal ENC loss is a novel pathogenic mechanism of HSCR, and suggest that NO donor drugs may be useful in treating ischemia-related ENC death.

## Acknowledgements

We thank all the donors and their parents for their consent and participation in this study.

## Contributors

Yuxia Z. and Yan Z. conceived the study and designed the experiments. Yuxia Z., Yan Z., F.B., and H.X. supervised the study. Yuxia Z., D.X., and W.L. wrote the manuscript with significant input from A.M.L., F.B., and Yan Z. D.X. performed the scRNA-seq, ST-seq, developed all code and analyzed the sequencing data. W.L. performed *in vitro* and *in vivo* experiments with assistance from L.L., Weiyong Z., M.Y., and C.L. W.L. analyzed the data generated from *in vivo* and *in vitro* experiment. C.L. collected and analyzed the clinical information with the assistance from Weiyong Z. and X.Z. Wenhuan Z., Q.W., Wei Z., and J.Z. recruited and provided patient care and clinical and histological assessments. All authors discussed and approved the manuscript.

## Data availability statement

The sequencing data involved in this study were deposited at the Genome Sequence Archive of Beijing Institute of Genomics, Chinese Academy of Sciences (GSA-Human accession: HRA006505). Data acquisition follows the guidelines of Genome Sequence Archive (http://bigd.big.ac.cn/gsa-human). All codes and resource data were available upon reasonable request to the authors

## References

1. Rao M, Gershon MD. The bowel and beyond: the enteric nervous system in neurological disorders. Nat Rev Gastroenterol Hepatol 2016;13(9):517–28. doi: 10.1038/nrgastro.2016.107 [published Online First: 2016/07/21]

2. Browning KN, Verheijden S, Boeckxstaens GE. The Vagus Nerve in Appetite Regulation, Mood, and Intestinal Inflammation. Gastroenterology 2017;152(4):730–44. doi: 10.1053/j.gastro.2016.10.046 [published Online First: 20161215]

3. Boesmans W, Nash A, Tasnady KR, et al. Development, Diversity, and Neurogenic Capacity of Enteric Glia. Front Cell Dev Biol 2021;9:775102. doi: 10.3389/fcell.2021.775102 [published Online First: 20220117]

4. Montalva L, Cheng LS, Kapur R, et al. Hirschsprung disease. Nat Rev Dis Primers 2023;9(1):54. doi: 10.1038/s41572-023-00465-y [published Online First: 2023/10/13]

5. Amiel J, Sproat-Emison E, Garcia-Barcelo M, et al. Hirschsprung disease, associated syndromes and genetics: a review. J Med Genet 2008;45(1):1–14. doi: 10.1136/jmg.2007.053959 [published Online First: 2007/10/30]

6. Gabriel SB, Salomon R, Pelet A, et al. Segregation at three loci explains familial and population risk in Hirschsprung disease. Nat Genet 2002;31(1):89–93. doi: 10.1038/ng868 [published Online First: 20020415]

7. Chatterjee S, Karasaki KM, Fries LE, et al. A multi-enhancer RET regulatory code is disrupted in Hirschsprung disease. Genome Res 2021;31(12):2199–208. doi: 10.1101/gr.275667.121 [published Online First: 20211115]

8. Uribe RA, Hong SS, Bronner ME. Retinoic acid temporally orchestrates colonization of the gut by vagal neural crest cells. Dev Biol 2018;433(1):17–32. doi: 10.1016/j.ydbio.2017.10.021 [published Online First: 20171103]

9. Frith TJR, Gogolou A, Hackland JOS, et al. Retinoic Acid Accelerates the Specification of Enteric Neural Progenitors from In-Vitro-Derived Neural Crest. Stem Cell Reports 2020;15(3):557–65. doi: 10.1016/j.stemcr.2020.07.024 [published Online First: 2020/08/29]

10. Vincent E, Chatterjee S, Cannon GH, et al. Ret deficiency decreases neural crest progenitor proliferation and restricts fate potential during enteric nervous system development. Proc Natl Acad Sci U S A 2023;120(34):e2211986120. doi: 10.1073/pnas.2211986120 [published Online First: 2023/08/16]

11. Monforte-Munoz H, Gonzalez-Gomez I, Rowland JM, et al. Increased submucosal nerve trunk caliber in aganglionosis: a “positive” and objective finding in suction biopsies and segmental resections in Hirschsprung’s disease. Arch Pathol Lab Med 1998;122(8):721–5.

12. Tani G, Tomuschat C, O’Donnell AM, et al. Increased population of immature enteric glial cells in the resected proximal ganglionic bowel of Hirschsprung’s disease patients. J Surg Res 2017;218:150–55. doi: 10.1016/j.jss.2017.05.062 [published Online First: 2017/10/08]

13. Zhou Y, Yang J, Watkins DJ, et al. Enteric nervous system abnormalities are present in human necrotizing enterocolitis: potential neurotransplantation therapy. Stem Cell Res Ther 2013;4(6):157. doi: 10.1186/scrt387 [published Online First: 2014/01/16]

14. Vasina V, Barbara G, Talamonti L, et al. Enteric neuroplasticity evoked by inflammation. Auton Neurosci 2006;126–127:264-72. doi: 10.1016/j.autneu.2006.02.025 [published Online First: 2006/04/21]

15. Holland AM, Bon-Frauches AC, Keszthelyi D, et al. The enteric nervous system in gastrointestinal disease etiology. Cell Mol Life Sci 2021;78(10):4713–33. doi: 10.1007/s00018-021-03812-y [published Online First: 2021/03/27]

16. Khoury-Hanold W, Yordy B, Kong P, et al. Viral Spread to Enteric Neurons Links Genital HSV-1 Infection to Toxic Megacolon and Lethality. Cell Host Microbe 2016;19(6):788–99. doi: 10.1016/j.chom.2016.05.008 [published Online First: 2016/06/10]

17. Elmentaite R, Kumasaka N, Roberts K, et al. Cells of the human intestinal tract mapped across space and time. Nature 2021;597(7875):250-55. doi: 10.1038/s41586-021-03852-1 [published Online First: 2021/09/10]

18. Li Z, Lui KN, Lau ST, et al. Transcriptomics of Hirschsprung disease patient-derived enteric neural crest cells reveals a role for oxidative phosphorylation. Nat Commun 2023;14(1):2157. doi: 10.1038/s41467-023-37928-5 [published Online First: 2023/04/16]

19. Fawkner-Corbett D, Antanaviciute A, Parikh K, et al. Spatiotemporal analysis of human intestinal development at single-cell resolution. Cell 2021 doi: 10.1016/j.cell.2020.12.016 [published Online First: 2021/01/07]

20. Elmentaite R, Ross ADB, Roberts K, et al. Single-Cell Sequencing of Developing Human Gut Reveals Transcriptional Links to Childhood Crohn’s Disease. Dev Cell 2020;55(6):771–83 e5. doi: 10.1016/j.devcel.2020.11.010 [published Online First: 2020/12/09]

21. Polanski K, Young MD, Miao Z, et al. BBKNN: fast batch alignment of single cell transcriptomes. Bioinformatics 2020;36(3):964–65. doi: 10.1093/bioinformatics/btz625 [published Online First: 2019/08/11]

22. Wolf FA, Angerer P, Theis FJ. SCANPY: large-scale single-cell gene expression data analysis. Genome Biol 2018;19(1):15. doi: 10.1186/s13059-017-1382-0 [published Online First: 2018/02/08]

23. Trapnell C, Cacchiarelli D, Grimsby J, et al. The dynamics and regulators of cell fate decisions are revealed by pseudotemporal ordering of single cells. Nat Biotechnol 2014;32(4):381–86. doi: 10.1038/nbt.2859 [published Online First: 2014/03/25]

24. Qiu X, Mao Q, Tang Y, et al. Reversed graph embedding resolves complex single-cell trajectories. Nat Methods 2017;14(10):979–82. doi: 10.1038/nmeth.4402 [published Online First: 2017/08/22]

25. Jin S, Guerrero-Juarez CF, Zhang L, et al. Inference and analysis of cell-cell communication using CellChat. Nat Commun 2021;12(1):1088. doi: 10.1038/s41467-021-21246-9 [published Online First: 2021/02/19]

26. Hao Y, Hao S, Andersen-Nissen E, et al. Integrated analysis of multimodal single-cell data. Cell 2021;184(13):3573–87 e29. doi: 10.1016/j.cell.2021.04.048 [published Online First: 2021/06/02]

27. Chen S, Zhou Y, Chen Y, et al. fastp: an ultra-fast all-in-one FASTQ preprocessor. Bioinformatics 2018;34(17):i884–i90. doi: 10.1093/bioinformatics/bty560 [published Online First: 2018/11/14]

28. Dobin A, Davis CA, Schlesinger F, et al. STAR: ultrafast universal RNA-seq aligner. Bioinformatics 2013;29(1):15–21. doi: 10.1093/bioinformatics/bts635 [published Online First: 20121025]

29. Newman AM, Steen CB, Liu CL, et al. Determining cell type abundance and expression from bulk tissues with digital cytometry. Nat Biotechnol 2019;37(7):773–82. doi: 10.1038/s41587-019-0114-2 [published Online First: 2019/05/08]

30. Zhou Y, Zhou B, Pache L, et al. Metascape provides a biologist-oriented resource for the analysis of systems-level datasets. Nat Commun 2019;10(1):1523. doi: 10.1038/s41467-019-09234-6 [published Online First: 2019/04/05]

31. Li H, Durbin R. Fast and accurate long-read alignment with Burrows-Wheeler transform. Bioinformatics 2010;26(5):589–95. doi: 10.1093/bioinformatics/btp698 [published Online First: 2010/01/19]

32. Danecek P, Bonfield JK, Liddle J, et al. Twelve years of SAMtools and BCFtools. Gigascience 2021;10(2) doi: 10.1093/gigascience/giab008

33. Institute B. Picard Toolkit: GitHub Repository; 2019 [Available from: https://broadinstitute.github.io/picard/.

34. McKenna A, Hanna M, Banks E, et al. The Genome Analysis Toolkit: a MapReduce framework for analyzing next-generation DNA sequencing data. Genome Res 2010;20(9):1297–303. doi: 10.1101/gr.107524.110 [published Online First: 2010/07/21]

35. Wang K, Li M, Hakonarson H. ANNOVAR: functional annotation of genetic variants from high-throughput sequencing data. Nucleic Acids Res 2010;38(16):e164. doi: 10.1093/nar/gkq603 [published Online First: 2010/07/06]

36. McLaren W, Gil L, Hunt SE, et al. The Ensembl Variant Effect Predictor. Genome Biol 2016;17(1):122. doi: 10.1186/s13059-016-0974-4 [published Online First: 2016/06/09]

37. Drokhlyansky E, Smillie CS, Van Wittenberghe N, et al. The Human and Mouse Enteric Nervous System at Single-Cell Resolution. Cell 2020;182(6):1606–22 e23. doi: 10.1016/j.cell.2020.08.003 [published Online First: 2020/09/06]

38. Demehri FR, Halaweish IF, Coran AG, et al. Hirschsprung-associated enterocolitis: pathogenesis, treatment and prevention. Pediatr Surg Int 2013;29(9):873–81. doi: 10.1007/s00383-013-3353-1 [published Online First: 2013/08/06]

39. Kuppe C, Ramirez Flores RO, Li Z, et al. Spatial multi-omic map of human myocardial infarction. Nature 2022;608(7924):766-77. doi: 10.1038/s41586-022-05060-x [published Online First: 20220810]

40. Ziegler M, Wang X, Peter K. Platelets in cardiac ischaemia/reperfusion injury: a promising therapeutic target. Cardiovasc Res 2019;115(7):1178–88. doi: 10.1093/cvr/cvz070

41. Ahmed NS, Lopes-Pires M, Pugh N. Zinc: an endogenous and exogenous regulator of platelet function during hemostasis and thrombosis. Platelets 2021;32(7):880–87. doi: 10.1080/09537104.2020.1840540 [published Online First: 20201115]

42. McCarthy N, Manieri E, Storm EE, et al. Distinct Mesenchymal Cell Populations Generate the Essential Intestinal BMP Signaling Gradient. Cell Stem Cell 2020;26(3):391–402 e5. doi: 10.1016/j.stem.2020.01.008 [published Online First: 20200220]

43. Ganfornina MD, Do Carmo S, Martinez E, et al. ApoD, a glia-derived apolipoprotein, is required for peripheral nerve functional integrity and a timely response to injury. Glia 2010;58(11):1320–34. doi: 10.1002/glia.21010 [published Online First: 2010/07/08]

44. Sladojevic N, Stamatovic SM, Johnson AM, et al. Claudin-1-Dependent Destabilization of the Blood-Brain Barrier in Chronic Stroke. J Neurosci 2019;39(4):743–57. doi: 10.1523/JNEUROSCI.1432-18.2018 [published Online First: 20181130]

45. Kubo F, Takeichi M, Nakagawa S. Wnt2b inhibits differentiation of retinal progenitor cells in the absence of Notch activity by downregulating the expression of proneural genes. Development 2005;132(12):2759–70. doi: 10.1242/dev.01856 [published Online First: 20050518]

46. Kubo F, Takeichi M, Nakagawa S. Wnt2b controls retinal cell differentiation at the ciliary marginal zone. Development 2003;130(3):587–98. doi: 10.1242/dev.00244

47. Chalazonitis A, Kessler JA. Pleiotropic effects of the bone morphogenetic proteins on development of the enteric nervous system. Dev Neurobiol 2012;72(6):843–56. doi: 10.1002/dneu.22002 [published Online First: 2012/01/04]

48. Zhao Y, Guan YF, Zhou XM, et al. Regenerative Neurogenesis After Ischemic Stroke Promoted by Nicotinamide Phosphoribosyltransferase-Nicotinamide Adenine Dinucleotide Cascade. Stroke 2015;46(7):1966–74. doi: 10.1161/STROKEAHA.115.009216 [published Online First: 20150609]

49. Kobayashi H, Yamataka A, Fujimoto T, et al. Mast cells and gut nerve development: implications for Hirschsprung’s disease and intestinal neuronal dysplasia. J Pediatr Surg 1999;34(4):543–8. doi: 10.1016/s0022-3468(99)90069-6 [published Online First: 1999/05/11]

50. Liu W, Zhang L, Wu R. Enteric Neural Stem Cells Expressing Insulin-Like Growth Factor 1: A Novel Cellular Therapy for Hirschsprung’s Disease in Mouse Model. DNA Cell Biol 2018;37(7):642–48. doi: 10.1089/dna.2017.4060 [published Online First: 2018/05/25]

51. Li S, Nie EH, Yin Y, et al. GDF10 is a signal for axonal sprouting and functional recovery after stroke. Nat Neurosci 2015;18(12):1737–45. doi: 10.1038/nn.4146 [published Online First: 2015/10/27]

52. Huang JY, Lynn Miskus M, Lu HC. FGF-FGFR Mediates the Activity-Dependent Dendritogenesis of Layer IV Neurons during Barrel Formation. J Neurosci 2017;37(50):12094–105. doi: 10.1523/JNEUROSCI.1174-17.2017 [published Online First: 2017/11/04]

53. Southard-Smith EM, Kos L, Pavan WJ. Sox10 mutation disrupts neural crest development in Dom Hirschsprung mouse model. Nat Genet 1998;18(1):60–4. doi: 10.1038/ng0198-60

54. Cummins EP, Crean D. Hypoxia and inflammatory bowel disease. Microbes Infect 2017;19(3):210-21. doi: 10.1016/j.micinf.2016.09.004 [published Online First: 2016/09/25]

55. Van Welden S, Selfridge AC, Hindryckx P. Intestinal hypoxia and hypoxia-induced signalling as therapeutic targets for IBD. Nat Rev Gastroenterol Hepatol 2017;14(10):596–611. doi: 10.1038/nrgastro.2017.101 [published Online First: 2017/08/31]

56. Giannotta M, Tapete G, Emmi G, et al. Thrombosis in inflammatory bowel diseases: what’s the link? Thromb J 2015;13:14. doi: 10.1186/s12959-015-0044-2 [published Online First: 2015/04/14]

57. Zezos P, Kouklakis G, Saibil F. Inflammatory bowel disease and thromboembolism. World J Gastroenterol 2014;20(38):13863-78. doi: 10.3748/wjg.v20.i38.13863 [published Online First: 2014/10/17]

58. Ji SG, Medvedeva YV, Wang HL, et al. Mitochondrial Zn(2+) Accumulation: A Potential Trigger of Hippocampal Ischemic Injury. Neuroscientist 2019;25(2):126–38. doi: 10.1177/1073858418772548 [published Online First: 2018/05/11]

59. Qi Z, Shi W, Zhao Y, et al. Zinc accumulation in mitochondria promotes ischemia-induced BBB disruption through Drp1-dependent mitochondria fission. Toxicol Appl Pharmacol 2019;377:114601. doi: 10.1016/j.taap.2019.114601 [published Online First: 2019/06/04]

60. Brown TA, Clayton DA. Release of replication termination controls mitochondrial DNA copy number after depletion with 2’,3’-dideoxycytidine. Nucleic Acids Res 2002;30(9):2004–10. doi: 10.1093/nar/30.9.2004

61. Lundberg JO, Weitzberg E. Nitric oxide signaling in health and disease. Cell 2022;185(16):2853–78. doi: 10.1016/j.cell.2022.06.010 [published Online First: 2022/08/06]

62. Lin M, Xian H, Chen Z, et al. MCM8-mediated mitophagy protects vascular health in response to nitric oxide signaling in a mouse model of Kawasaki disease. Nature Cardiovascular Research 2023;2(8):778–92. doi: 10.1038/s44161-023-00314-x

63. Nisoli E, Falcone S, Tonello C, et al. Mitochondrial biogenesis by NO yields functionally active mitochondria in mammals. Proc Natl Acad Sci U S A 2004;101(47):16507–12. doi: 10.1073/pnas.0405432101 [published Online First: 2004/11/17]

64. Fadista J, Lund M, Skotte L, et al. Genome-wide association study of Hirschsprung disease detects a novel low-frequency variant at the RET locus. Eur J Hum Genet 2018;26(4):561–69. doi: 10.1038/s41431-017-0053-7 [published Online First: 2018/01/31]

65. Tang CS, Gui H, Kapoor A, et al. Trans-ethnic meta-analysis of genome-wide association studies for Hirschsprung disease. Hum Mol Genet 2016;25(23):5265–75. doi: 10.1093/hmg/ddw333 [published Online First: 2016/10/06]

66. Bethell G, Wilkinson D, Fawkner-Corbett D, et al. Enteric nervous system stem cells associated with thickened extrinsic fibers in short segment aganglionic Hirschsprung’s disease gut are absent in the total colonic and intestinal variants of disease. J Pediatr Surg 2016;51(10):1581–4. doi: 10.1016/j.jpedsurg.2016.06.006 [published Online First: 20160616]

67. Moore SW. Total colonic aganglionosis and Hirschsprung’s disease: a review. Pediatr Surg Int 2015;31(1):1–9. doi: 10.1007/s00383-014-3634-3 [published Online First: 20141031]

68. Jaffe T, Thompson WM. Large-Bowel Obstruction in the Adult: Classic Radiographic and CT Findings, Etiology, and Mimics. Radiology 2015;275(3):651–63. doi: 10.1148/radiol.2015140916

