## supplemental method and figures for "Ischemia promotes hypertrophic nerve trunk formation and enteric neuron cell death in Hirschsprung’s disease"

**Supplementary Material**

**Supplementary Methods**

**Whole-exome library preparation and sequencing**

QIAamp DNA Mini Kit (Qiagen, Cat# 51304) were used to extract DNA from blood. In brief, blood was lysed in AL solution containing proteinase K; then the DNA was bound to the QIAamp Mini spin column, and was washed with buffer AW1 and buffer AW2; purified DNA was dissolved in DEPC water and stored at -80 ℃.

Purified DNA were processed using NEBNext® Ultra™ II DNA Library Prep Kit for Illumina® (NEB, Cat# E7645L) and SureSelectXT Human All Exon V6 (Agilent Technologies, Cat# 54898) according to the manufacturer’s protocol. Briefly, genomic DNA fragmented into 250 bp using Covaris system (Covaris); then the fragmented DNA was purified by Agencourt AMPure XP SPRI beads (Beckman Coulter, Cat# A63882); the ends of the purified DNA were repaired and were added with the adapters; the DNA fragments were further enriched by PCR after another purification using Agencourt AMPure XP SPRI beads; the products with exon regions were hybridized and amplified. The final products were quality-checked and sequenced with Illumina NovaSeq 6000 to generated 2 × 150 bp PE reads.

**Exome sequencing data analysis**

The sequencing data was first filtered using fastp^1^ (v.0.23.2) with adjusted parameters (-q = 19, -l = 36, -u = 50, others = default). A pipeline modified from "GATK Best Practices" were used to detect rare variants for HSCR donors. In brief, bwa^2^ (v.0.7.17) was used to align reads to the human reference genome (GRCh38); the outputs were converted to BAM (binary alignment map) files using samtools^3^ (v.1.13); picard^4^ (v.2.21.4) was used to sort the BAM files and mark the duplicated reads; GATK^5^ (v.4.2.0.0) with default parameters was used to process the sorted reads and generate VCF files containing variants for each donor.

The VCF files were annotated by VEP^6^ (v.104). Variants with a total allele frequency more than 1% were discarded. The results of VEP were further annotated with ANNOVAR^7^ (v.2020-06-08).

**Bulk transcriptome library preparation and sequencing**

FreeZol Reagent kit (Vazyme, Cat# R711-01) was used to extract the total RNA from the tissue according to the manufacturer’s protocol. In brief, colonic tissues were frozen in liquid nitrogen and grounded into powder; FreeZol Reagent was then added to the powder; the mixture was pipetted until fully lysed, and was incubated at room temperature for 5 minutes (min); dilution buffer was added to the lysis system, mixed with shaking, and incubated at room temperature for 5 min; the mixture was centrifuged at 12,000 × g at room temperature for 15 min; the supernatant was transferred to a new centrifuge tube; an equal volume of isopropyl alcohol was added to the centrifuge tube; the mixture was incubated at room temperature for 10 min and was centrifuged at 12,000 × g for 10 min at room temperature; the supernatant was discarded and the pellets were mixed with 1 mL of 75% ethanol; centrifuge at 8,000 × g for 3 min at room temperature, discard the supernatant, and keep the pellets; repeat the previous two steps once; dry the precipitate at room temperature; dissolve the precipitate in DEPC water. The total RNA was quantified and quality-checked by Qubit™ 4 Fluorometer (Invitrogen) and Agilent 2100 (Agilent Technologies).

The cDNA library was constructed using the NEBNext Ultra II RNA Library Prep Kit for Illumina (New England Biolabs Inc, Cat# E7775) according to manufacturer's instructions. In brief, mRNA was enriched from 1 μg total RNA using magnetic beads; the mRNA was fragmented to about 200 bp by Covaris system; the fragmented products were amplified to cDNA using random oligonucleotide primers; Sample Purification Beads were used to purify the cDNA; the purified products were end-repaired and the adapters were added; the product was purified by Sample Purification Beads again; the product were amplified with indexes by PCR, and was purified by Sample Purification Beads. The final products were quality-checked and sequenced with Illumina Novaseq 6000 to generated 2 × 150 bp PE reads.

The sequenced reads were filtered with fastp (v.0.23.2) with adjusted parameters (-q = 19, -l = 36, -u = 50, others = default). STAR (v.2.7.4a) with adjusted parameters (--readFilesCommand = gunzip, --quantMode = GeneCounts, others = default) was used to align reads to human genomics (GRCh38) and generate the count of gene expression. The *AddModuleScore* of Seurat package^8^ (v.4.0.1) was used to calculate the score of gene list (**Supplementary table 2**).

**Single-cell transcriptome library preparation and sequencing**

The colonic tissue was soaked in 4 ℃ DMEM (GIBCO, Cat# 11966025) and was processed immediately when was transported to the laboratory: the tissue was cut into pieces on ice with ophthalmic scissors; fragments of tissues were incubated with 1 mg/mL collagenase 1A (Sigma, Cat# C2674-500mg) and 10 U/mL DNase I (Roche, Cat# 11284932001) for 35 min on a 37°C shaker (50 r/min); suspension filtered by a 70 μm filter was centrifuged at 700 × g for 5 min at 4°C; cells were resuspended with 1 × PBS (HyClone, Cat# SH30256.01) and stained with propidium iodide (Invigentech, Cat# PI0100) for 5 min on ice; flow cytometry (FACSAria SORP flow cytometer, BD science) was used to enrich live cells.

Chromium Next GEM Single Cell 5' Kit v2 (10x Genomics, Cat# 1000263) were used to construct the cDNA library following manufacturer's instructions: single-cell suspension, RT-PCR master mix, and nanoliter-scale gel beads were mixed and were loaded to single-cell chip; nanoliter-scale water-in-oil system was formed, cells were lysed, mRNAs were captured and were reverse transcribed into cDNA library containing cell barcodes. Quality-checked library was sequenced with Illumina Novaseq 6000.

**Prerocessing scRNA-seq data**

Cell Ranger (v.6.1.1) with default parameters was used to align reads to human reference genome (refdata-gex-GRCh38-2020-A) and quantify gene expression. The outputs were processed with scanpy package^9^ (v.1.7.1) following official tutorials: raw count matrix, gene table, and barcodes table of each sample were loaded; cells with fewer than 200 genes were discarded; cells with mitochondrial transcripts accounting for more than 30% were eliminated; genes expressed in less than 3 cells were discarded; filtered matrixes of each sample were merged to form an integrated matrix; the raw counts were normalized, log-transformed, and scaled; the dimensionality of the data was reduced by principal component analysis; then BBKNN^10^ was run to remove the batch effect and to compute the neighborhood graph; the neighborhood graph was embedded into two dimensions using UMAP; Leiden graph-clustering method was used to cluster the neighborhood graph. The rank of differentially expressed genes (DEGs) of single-cell clusters were computed by two-side Mann-Whitney-U test.

**Annotation of single-cell clusters**

Single-cell clusters were first divided into 8 lineages (B cells: *PTPRC^+^ CD19^+^ / IGHA^+^*; T and innate lymphoid cells: *PTPRC^+^ CD3E^+^*; myeloid cells: *PTPRC^+^ CD3E^-^ CD19^-^*; fibroblast-like cells: *PTPRC*^-^ *RGS5^-^* Collagen*^+^*; perivascular cells: *PTPRC*^-^ *RGS5*^+^; endothelial cells: *PTPRC*^-^ *PECAM1*^+^; epithelial cells: *PTPRC*^-^ *EPCAM*^+^; glial cells: *PTPRC*^-^ *S100B*^+^). Complete cell annotations were summarized in **Supplementary Table 4**.

**Prediction of cell differentiation trajectory**

Monocle 3^11, 12^ (v.1.0.0) was used to predict cell differentiation trajectory base on the results of UMAP. The process was run with default parameters. Genes differential expressed during differentiation (*p*-adj < 0.05, Moran's I > 0.1) were used for pathway enrichment.

**Prediction of cell-cell interaction**

A raw sub-matrix was first randomly sampled (500 cells for each single-cell cluster) and processed with Seurat^8^ (v.4.0.1). *SCTransform* of Seurat were used to normalize the matrix, other functions were run with default parameters. CellChat^13^ (v.1.1.0) with default parameters was used for predicting cell-cell communication between single-cell populations. Ligand-receptor pairs occurred in less than 10 cells were discarded.

**Spatial transcriptome library preparation and sequencing**

The colonic tissue was transported to laboratory in 4 ℃ DMEM and was processed immediately according to the following procedures: tissue was trimmed to appropriate size (about 6 × 6 × 10 mm) and was quick-frozen in isopentane at -196 ℃; the frozen tissue was embedded with 4 ℃ optimal cutting temperature compound (Tissue-Tek, Cat# SAKURA Tissue 4583) and the whole mixture was frozen at -80 ℃ immediately.

Visium Spatial Tissue Optimization Slide & Reagents Kit (10x Genomics, Cat# 1000193) and Visium Spatial Gene Expression Slide & Reagents Kit (10x Genomics, Cat# 1000187) were used for constructing spatial transcriptome library following the manufacturer's instructions: sample with RNA integrity number larger than 7 were used for library building; tissues were cut into 10 μm thick slices; H&E staining were used to visualize the histological structure; by lysing the tissue, the mRNA is released and captured by primers containing spatial information on the chip; after reverse transcription and amplification, cDNA library with spatial information was built and sequenced with Illumina Novaseq 6000.

**Processing ST-seq data**

Spaceranger (v.1.3.1) was used to process the spatial sequencing data according to official tutorials: raw reads were aligned to human reference genome (refdata-gex-GRCh38-2020-A); then, the expression of genes was quantified and split into spatial spots; finally, the gene expression was linked with histological structures.

The Seurat package^8^ (v.4.0.1) was used for processing the outputs of Spaceranger based on official tutorials: *sctransform* were used for data normalization; *merge* was used to generate an integrated object containing all slides; PCA dimensionality reduction was run to reveal the main axes of variation; then 20.param nearest neighbors was computed to construct a shared nearest neighbor graph; ST clusters were identified by running *FindClusters* with defaulted parameters; the shared nearest neighbor graph was further embedded into two dimensions using UMAP.

A semi-supervised method was used for annotating ST clusters. ST clusters were first generated in an unsupervised way. C1 and C2 located in the mucosal layer. C3-C7 located in the submucosal layer. C11 located in the large blood vessels. C12 and C13 located in the nerve-related regions. There were various muscular ST clusters. Except for C10, most muscular ST clusters were sample-specific and were annotated base on their location (circular muscular ST cluster: C8; longitudinal muscular ST cluster: C9). C10, as a group of cross-sample muscular ST spots that only occurred in ganglionic HSCR colons, was defined as a unique ST cluster. *FindMarkers* in Seurat^8^ (v.4.0.1) was used to identified DEGs for processed ST clusters based on two-side Mann-Whitney-U test. Given that none upregulated DEGs were identified in C6 and C6 located in blank areas, C6 was identified as low-quality ST clusters and was excluded from following analysis.

**Predicting the abundance of single-cell clusters in ST spots**

Seurat package^8^ (v.4.0.1) were used to link cell abundance to spatial distribution: a randomly sampled single-cell Seurat object containing 500 cells for each single-cell cluster were generated according to abovementioned way; *FindTransferAnchors* was used to find a set of anchors between single-cell reference and ST-seq data; *TransferData* was used to link the cell abundance to the ST spot.

**Imputing the abundance of ST clusters in bulk RNA-seq data**

CIBERSORTx^14^ (https://cibersortx.stanford.edu/) was used to estimate the abundance of ST clusters in bulk RNA-seq data. The single-cell reference was the raw count matrix of the randomly sampled single-cell Seurat object mentioned above. S-mode was applied to eliminate the batch effect.

**Fluorescent staining and visualization**

Formalin-fixed paraffin-embedding (FFPE) sections of surgical samples were processed as reported previously. In brief, sections were dewaxed to water and processed with blocking buffer (PBS with 5% normal donkey or goat serum and 0.3% Triton X-100) for 1 hour (hr) at room temperature. Then, sections were stained with primary antibodies in a wet chamber at 4 ℃ overnight, followed by PBS washing. After that, sections were incubated with secondary antibodies for an hr at room temperature, then were mount with VECTASHIELD Antifade Mounting Medium with DAPI (Vector Laboratories, Cat# H-1200). All staining process were done in a dark condition.

**Immunohistochemical staining**

Deparaffinized tissue sections were subjected to antigens rederivation, peroxidase removal, 3% BSA blockage and then were incubated with indicated primary antibodies at 4°C overnight followed by appropriate secondary HRP-conjugated antibodies before staining with 3,3'-diaminobenzidine Substrate Kit (Pierce, Cat#34002). Nuclei were counterstained with hematoxylin. Sections incubated with species-matched IgG alone were used as negative controls.

**Masson's trichrome staining**

Masson's trichrome staining (Servicebio, Cat# G1006-100ML) was performed following the Manufacturer's instructions: FFPE sections were dewaxed to water and incubated in Masson A buffer at room temperature for 15 hr; then the mixture was incubated at 65 ℃ for 30 min; the sections were rinsed with pure water and were incubated in B/C (1 : 1) mixed buffer for 1 min; the sections were immersed in 1% hydrochloric acid ethanol solution; Buffer D (65 ℃, 6 min), E (room temperature , 1 min), and F (65 ℃, 30 s) were then used to incubate the sections; the sections were then rinsed with 1% glacial acetic acid for 8 s three times; then the sections were dehydrated with ethanol, n-butanol, and xylene.

**Flow cytometry detecting cell death and mitochondria damage**

Stimulated SH-SY5Y cells were collected and stained with AnnexinV-FITC and PI (Invigentech, Cat# PI0100) for 20 min and then detected by BD Aria (BD Biosciences, CA, USA).

Stimulated SH-SY5Y cells were stained with MitoTracker Green FM (300 nM, 30 min) (Thermo Fisher Scientific, Cat# M7512), FluoZin-3 (10 μM, 30 min) (Invitrogen, Cat# F24195) or TMRM (100 nM, 30 min) (Cell Signaling, Cat# 92043S). Cells were washed with PBS and acquired by BD Aria.

**Reverse transcription and real-time PCR**

Fresh tissues or cells were submerged in RNA*later* (Thermo Fisher Scientific, Cat# 15596018) at 4°C overnight. Total RNA was extracted and purified by using the RNeasy Mini Kit (Qiagen, Cat#74104) according to the manufacturer’s protocol. Equal amounts (2 µg) of total RNA were reverse-transcribed to cDNA by using All-in-One RT MasterMix (Applied Biological Materials Inc., Cat# G592). After reverse transcription, real-time PCR amplification was performed using SYBR Green qPCR Mastermixes (Qiagen, Cat# 330513) under a 7500 Real-Time PCR System (Applied Biosystems, NY). PCR program consisted of 95 °C for 5 min; 40 cycles of 95°C for 30 seconds (s), 55°C for 30 s, and 72°C for 30 s; and 72°C for 5 min. The expression of candidate genes in each group of mice was normalized to *Hprt* to obtain a ΔCt value and to calculate a 2^-(mean ΔCt).

**Establishment of the neonatal ischemic enterocolitis (NEI) mouse model**

Neonatal ischemic enterocolitis was induced in 5-day-old mice of either gender, which were randomly divided into control and experimental groups, by gavage feeding newborn mice with enteric bacteria obtained from SPF wild type C57BL/6 mice or Sox10WT/MUT C3H mice once per day for the first and second day of the experiment. Additionally, the mice were subjected to hypoxia (5% O_2_-95% N_2_) for 2 min in a hypoxia chamber (STEMCELL) twice daily and followed by 4 °C treatment for 10 min from the third to fifth day. Mice were then sacrificed for histopathological analysis. Colon tissues were collected for further analyses.

**Transmission electron microscope**

Cells or tissues were rinsed and fixed in 2.5% glutaraldehyde buffer (Servicebio, Cat# G1102) at 4 °C overnight, followed by rinsing in sodium cacodylate, fixing in 1% osmium tetroxide for 1 h, staining in 2% uranyl acetate in maleate buffer (pH 5.2) for 1 h and rinsed and dehydrated in an ethanol series and infiltrated with resin (EMS) and incubated overnight at 60 °C. Hardened blocks were cut using a Leica Ultracut UCT. Then, 60-nm sections were collected on formvar/carbon-coated grids and contrast stained using 2% uranyl acetate and lead citrate.

**Calculating the index of clinical parameters**

The clinical data was first ranked from lowest to highest. Then the results of sorting were scaled for visualization.

**Supplementary Figures**

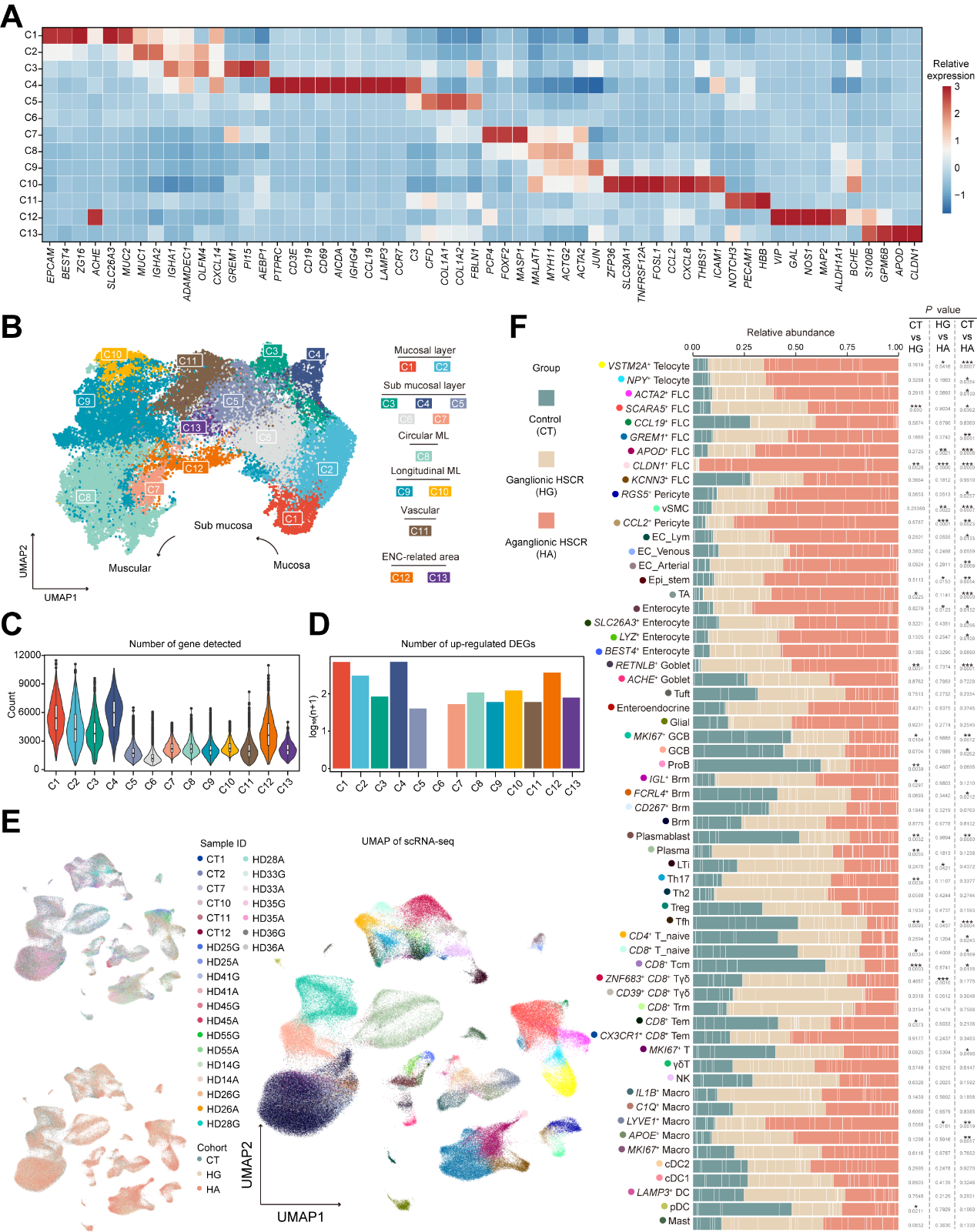
 **Supplemental figure 1.** scRNA-seq and ST-seq characteristics of control and HSCR colons.

(**A**) Heatmap showing expression of marker genes of ST clusters. Colors represent center-normalized relative expression levels.

(**B**) UMAP showing the annotation of 13 ST-clusters.

(**C**) Violin-box plot showing number of genes detected in ST clusters (box limit: standard deviation; line within the box: median; whiskers: 1.5 × outliers).

(**D**) Bar plot showing the number of up-regulated DEGs detected in ST clusters. n, the number of up-regulated DEGs.

(**E**) UMAP showing sample origination (left panel) and 61 scRNA subsets (right panel). The corresponding annotation of each subset was shown in **(supplemental figure 1F)**.

(**F**) Bar plot displaying group compositions of scRNA-seq sub-populations. Each block represents a subject. *P* values were calculated based on two-way ANOVA^15^. ****P* < 0.001, ***P* < 0.01, **P* < 0.05. FLC, fibroblast-like cell. vSMC, vascular smooth muscular cell. EC_Lym, lymphatic endothelial cell. EC_Venous, venous endothelial cell. EC_Arterial, arterial endothelial cell. Epi_stem, epithelial stem cell. TA, transit amplifying cell. GCB, germinal center B cell. ProB, B cell precursor cell. Brm, tissue resident memory B cell. LTi, lymphoid tissue inducer cell. Th17, helper T 17 cell. Th2, helper T 2 cell. Treg, regulatory T cell. Tfh, T follicular helper cell. T_naïve, naïve T cell. Tcm, central memory T cell. Tγδ, a mixture of γδ T cells and αβ T cells. Trm, tissue resident memory T cell. Tem, effector memory T cell. γδT, γδ T cell. NK, natural killer cell. Macro, macrophage. cDC, conventional dendritic cell. DC, dendritic cell. pDC, plasmacytoid dendritic cell.

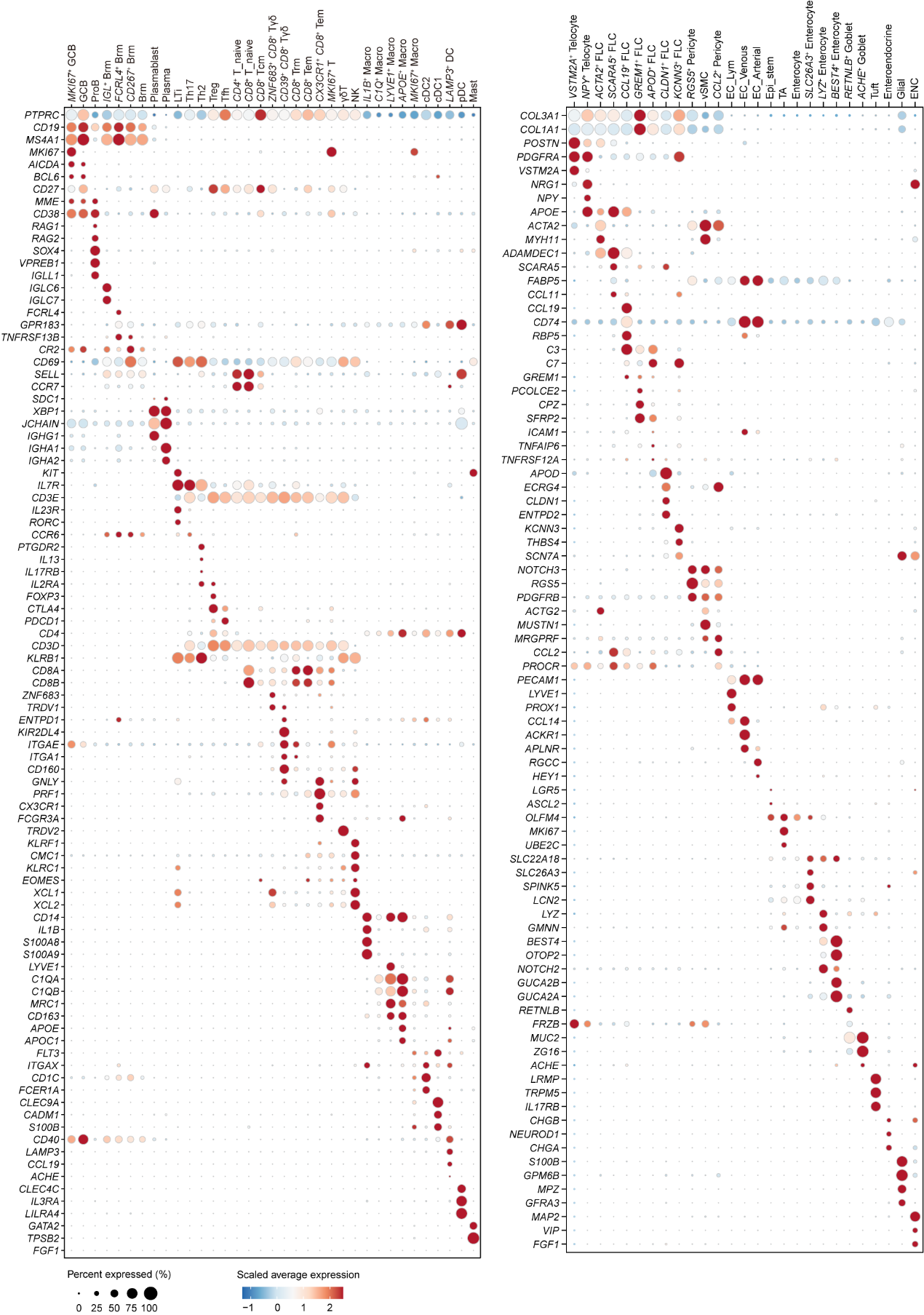

**Supplemental figure 2.** Expression of marker genes in single-cell reference.

Heatmap showing the expression of marker genes in 62 single-cell populations. Exogenous ENC data^16^ was included. The size of the dot indicates the proportion of cells expressing a specific gene in a single-cell subpopulation. The original expression count n is log_2_(n+1) transformed and averaged among the cells within the subpopulation. The color represents the mean expression scaled by row.

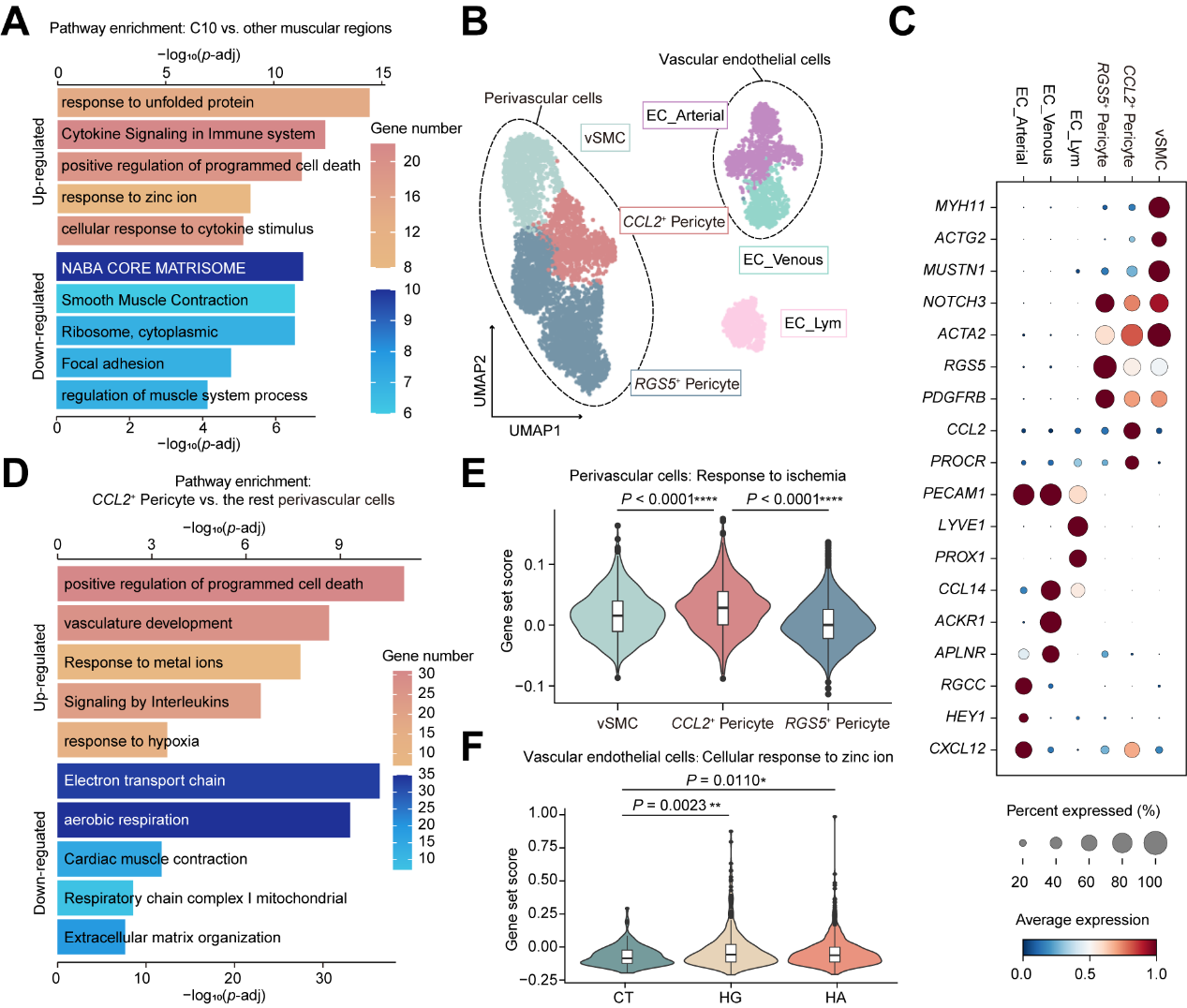

**Supplemental figure 3.** Remodeling of blood vessels in HSCR colons.

(**A**) Bar plot displaying the enriched pathways for the up- and down-regulated DEGs between C10 and the other muscular ST-clusters (C8 and C9).

(**B**) UMAP displaying the annotation of sub-populations of perivascular cells and endothelial cells captured by scRNA-seq.

(**C**) Heatmap showing the expression of marker genes in perivascular cells and endothelial cells captured by scRNA-seq. The expression of genes is scaled by row and presented by color. Dot size represents the percentage of cells expressing corresponding genes.

(**D**) Bar plot displaying enriched pathways for the up- and down-regulated DEGs between *CCL2*^+^ pericytes and other perivascular cells.

(**E**) Violin-box plot displaying the scores of ischemia response (GO: 0002931) gene set in perivascular cells captured by scRNA-seq.

(**F**) Violin-box plot showing the gene set score of zinc ion response (GO: 0071294) in vascular endothelial cells (EC_Arterial and EC_Venous) captured by scRNA-seq. Statistical analysis was performed using two-sided Mann-Whitney U test (**E-F**). **** *P* < 0.0001, ****P* < 0.001, ***P* < 0.01, **P* < 0.05. Data are median (line within the box) with a SD (box limit) and 1.5 × outliers (whiskers) (**E-F**).

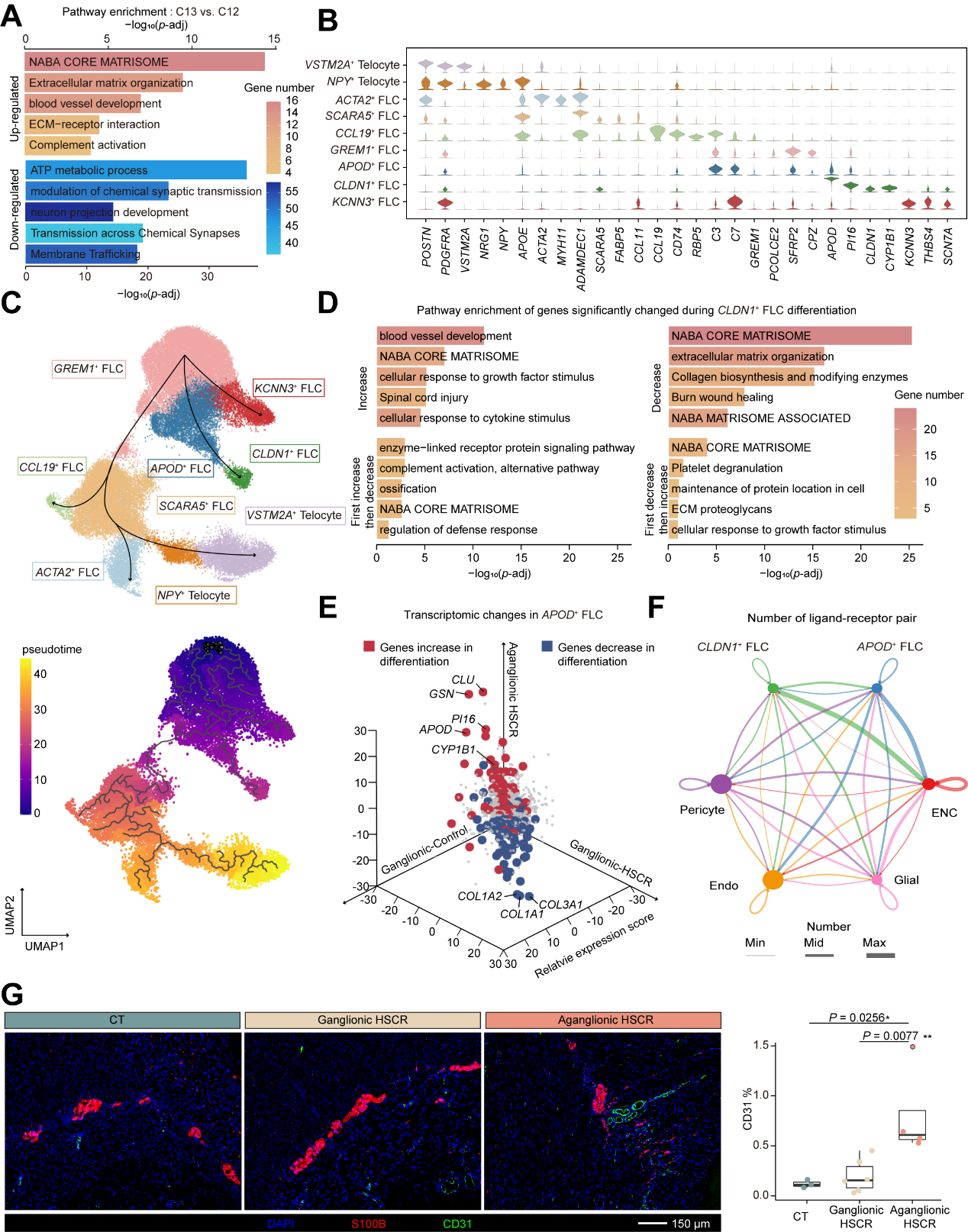

**Supplemental figure 4.** Remodeling of neural microenvironment in aganglionic HSCR colons.

(**A**) Bar plot displaying enriched pathway for the DEGs between ganglia (C12) and HNTs (C13).

(**B**) Violin plot showing the expression of marker genes for single-cell FLC populations.

(**C**) UMAP displaying the annotation (upper panel) and the pseudo time trajectory (lower panel) of FLCs populations.

(**D**) Bar plot showing the DEG-related pathways following the differentiation of *GREM1*^+^, *APOD*^+^ and *CLDN1*^+^ FLCs.

(**E**) 3D volcano plot displaying the transcriptional changes among *APOD*^+^ FLCs between control and HSCR subjects. Increased and decreased DEGs in the differentiation of *CLDN1*^+^ FLCs were marked with thick colored dots.

(**F**) Circle plot showing the number of ligand-receptor pairs identified by cell-cell interaction analysis. The thickness of the line represents the number of ligand-receptor pairs.

(**G**) IF imaging and box plot (box limit: standard deviation; line within the box: median; whiskers: 1.5 × outliers) showing the peri-ganglia and peri-HNTs vessels in colonic tissues (three CT, six HG and four HA). Each dot represents a donor. One-tailed *t*-test. ****P* < 0.001, ***P* < 0.01, **P* < 0.05.

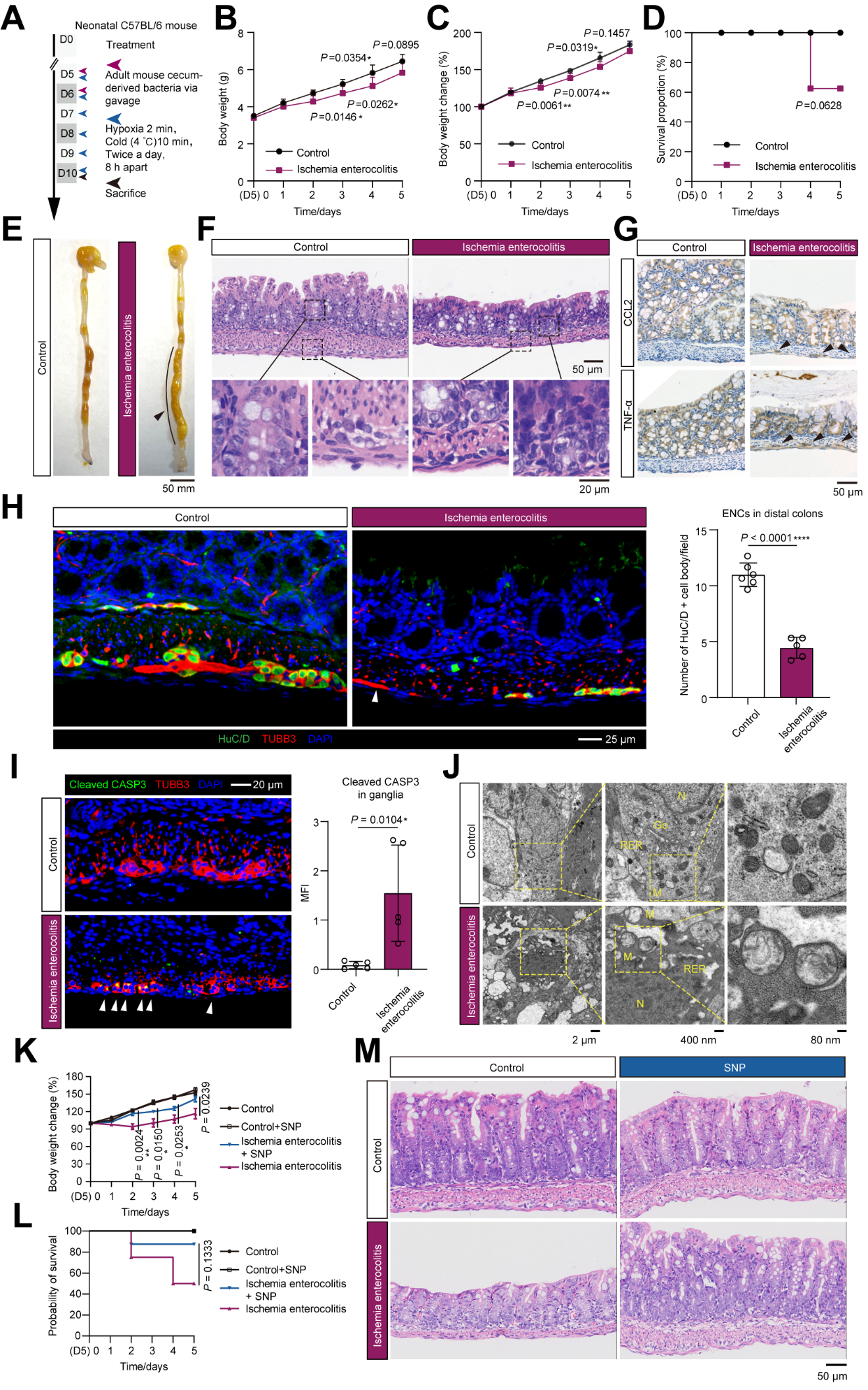

**Supplemental figure 5.** Characterization of the neonatal ischemic enterocolitis mouse model.

(**A**) Diagram showing the establishment of a neonatal ischemic enterocolitis (NIE) model.

(**B**) Line chart showing the changes in body weight of the mice.

(**C**) Line chart displaying the changes in body weight by percentage for the mice.

(**D**) Line chart showing the survival curve of the mice.

(**E**) Picture showing fecal retention in the NIE mice.

(**F**) H&E staining displaying the histological characteristics of the mouse colonic tissues.

(**G**) Immunohistochemistry (IHC) imaging showing TNF-α and CCL2 levels in the control and NIE mice. Ganglia are indicated by black arrows.

(**H**) IF imaging and bar plot displaying ganglia in the distal colons of the mice. TUBB3 (red) represents nerve fibers. HuC/D (green) represents nerve bodies. HNTs containing nerve fibers while lacking nerve bodies were indicated by white triangles.

(**I**) IF imaging and bar plot showing cleaved CASP3 in colonic ganglia in control and NIE mice.

(**J**) TEM displaying apoptosis-related changes of the ENCs in control and NIE mice. M, mitochondria. N, nucleus. RER, rough endoplasmic reticulum. Go, Golgi apparatus.

(**K**) Line char showing the effects of sodium nitroprusside on mouse body weight. SNP, sodium nitroprusside.

(**L**) Line char showing the effects of sodium nitroprusside on the survival of mice.

(**M**) H&E staining displaying the effects of sodium nitroprusside on the morphology of mouse colon tissues. Data were presented as mean ± SD (**B-D, H-I, K-L**). Each datapoint represents an animal (**H-I**). Statistical analysis was performed using two-tailed *t*-test (**B-C, H-I, K**) or log-rank test (**D**, **L**). *****P* < 0.0001, ****P* < 0.001, ***P* < 0.01, **P* < 0.05. Images and statistical results were generated from 5 (**B-G**, **I-M**) or 6 (**H**) control mice, 5 control mice injected with sodium nitroprusside (**K-M**), 5 NIE mice (**B-M**) and 5 (**K-M**) NIE mice injected with sodium nitroprusside.

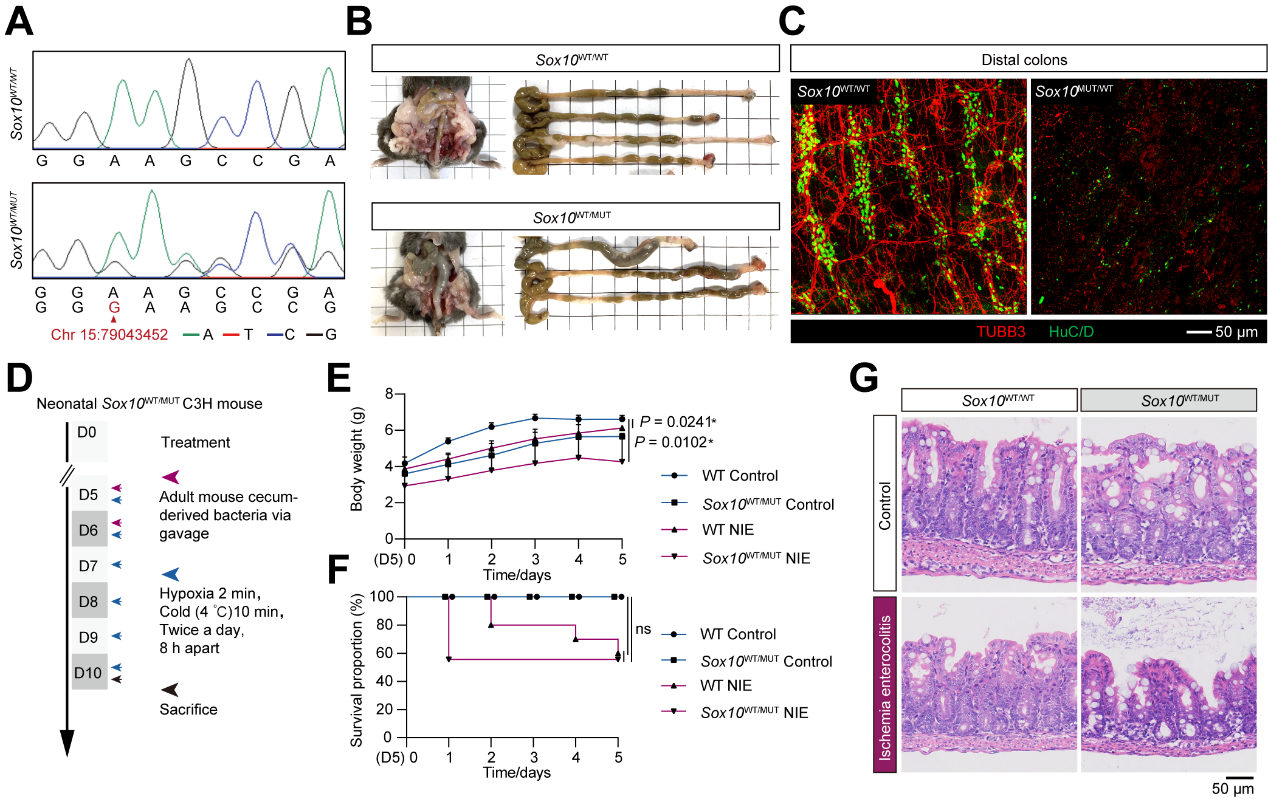

**Supplemental figure 6.** Characterization of *Sox10*^WT/MUT^ mice.

(**A**) Sanger sequencing data showing the genetic background WT and the *Sox10*^WT/MUT^ C3H mice.

(**B**) Pictures showing fecal retention in *Sox10*^WT/MUT^ mice.

(**C**) IF imaging displaying ENC-related structures in the distal colons of WT and *Sox10*^WT/MUT^ mice. TUBB3 (red) represents nerve fibers. HuC/D (green) represents nerve bodies.

(**D**) Diagram showing the establishment of the NIE model using *Sox10*^MUT/WT^ and WT neonatal mice.

(**E-F**) Line charts showing the changes of weight (**E**) and survival of mice (**F**).

(**G**) H&E images displaying histological characteristics of WT and *Sox10*^WT/MUT^ mice under untreated or NIE conditions. Representative images and data were generated from four WT animals and three *Sox10*^MUT/WT^ animals (**A-G**).

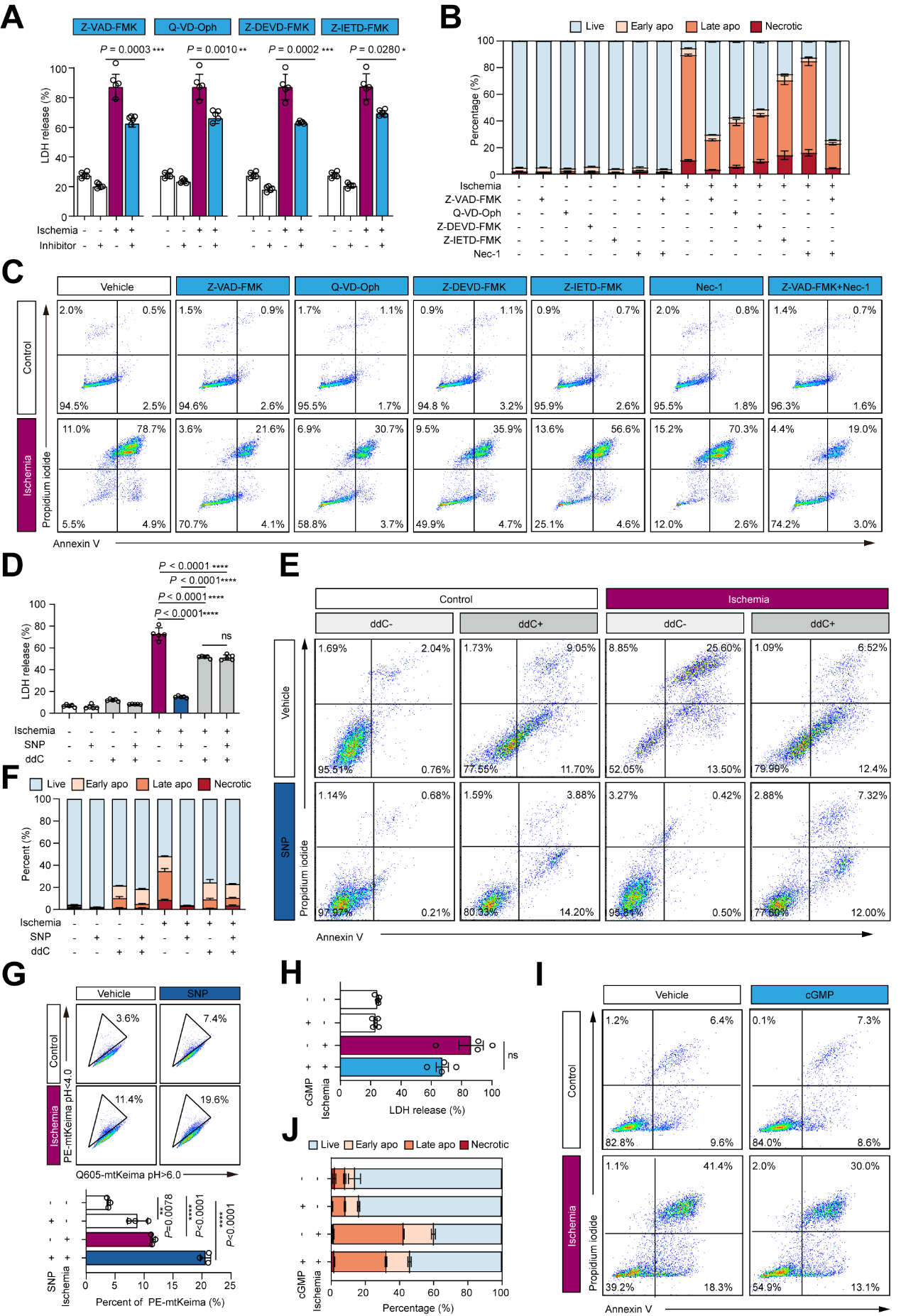

**Supplemental figure 7.** Damaged mitochondria mediate ischemia-related neuron apoptosis.

(**A**) Bar plot showing the impacts of CASP inhibitors on LDH release in SH-SY5Y cells under indicated treatments. Z-VAD-FMK (50 μM), a pan-caspase inhibitor. Q-VD-Oph (50 μM), a pan-caspase inhibitor. Z-DEVD-FMK (50 μM), a CASP8 inhibitor. Z-IETD-FMK (500 μM), a CASP3 inhibitor. LDH (Lactate dehydrogenase) release indicate cell death.

(**B-C**) Bar plot (**B**) and flow cytometry chart (**C**) displaying the impacts of CASP inhibitors and necroptosis inhibitor (Nec-1) on the survival of SH-SY5Y cells. Apo: apoptosis.

(**D**) Bar plot displaying the effects of mitochondrial removal on LDH release of SH-SY5Y cells under indicated conditions. ddC, an agent depletes mitochondria by inhibitng the mtDNA polymerase POLG.

(**E-F**) Flow cytometry chart (**E**) and bar plot (**F**) displaying the effects of mitochondrial removal on the survival of SH-SY5Y cells under different treatments.

(**G**) Flow cytometry chart and bar plot displaying mitophagy activities of SH-SY5Y cells under indicated conditions. mtKeima, a fluorescent protein indicating mitophagy at PH<4.

(**H**) Bar plot showing the impacts of cGMP signaling on the releasing of LDH in SH-SY5Y cells under different treatments.

(**I-J**) Flow cytometry chart (**I)** and bar plot (**J**) showing impacts of cGMP signaling on the survival of LDH in SH-SY5Y cells under different treatments. cGMP, 8-Bromo-cGMP sodium. Horizontal bars represent mean ± SD generated from three (**G**), four (**H**, **J**), or five (**A**, **B**, **D**, **F**) independent experiments. Each datapoint represents an independent experiment. Statistical analysis was performed using two-tail *t*-test (**A**, **D**, **G**, **H**, **J**). *****P* < 0.0001, ****P* < 0.001, ***P* < 0.01, **P* < 0.05. ns, *P* > 0.05.

**Supplemental table 1.** Clinical information of participants (see Supplementary Table 1.xlsx). The random ID (Masked_ID) cannot reveal the identity of the study subjects and was not known to anyone outside the research group.

**Supplemental table 2.** List of susceptible genes for HSCR and genes used for calculating gene set score (see Supplementary Table 2.xlsx).

**Supplemental table 3.** Rare variants detected in susceptible genes for HSCR (see Supplementary Table 3.xlsx).

**Supplemental table 4.** Annotation of single-cell subpopulations (see Supplementary Table 4.xlsx).

**Supplemental table 5. Key resources**

| REAGENT or RESOURCE | SOURCE | IDENTIFIER |
| --- | --- | --- |
| Antibodies | | |
| Rabbit monoclonal [EPRR18871] to Claudin 1 | abcam | Cat# ab211737 |
| Rabbit monoclonal [EP1576Y] to S100 beta | abcam | Cat# ab52642 |
| Rabbit monoclonal [EPR19691] to MAP2 | abcam | Cat# ab183830 |
| Rabbit polyclonal to Neuron specific beta III Tubulin | abcam | Cat# ab229590 |
| Rabbit polyclonal to Collagen III | abcam | Cat# ab7778 |
| Rabbit polyclonal to Metallothionein | abcam | Cat# ab192385 |
| Mouse monoclonal [C31.3] to CD31 | abcam | Cat# ab187377 |
| Rabbit monoclonal [EPR4330] to CD41 | abcam | Cat# ab134131 |
| Rabbit polyclonal to CD45 | abcam | Cat# ab10558 |
| Human CCL2/JE/MCP‑1 Antibody | biotechne | Cat# AF-279-NA |
| Cleaved Caspase-3 (Asp175) Antibody | CST | Cat# 9661 |
| Claudin 1 Monoclonal Antibody | Invitrogen | Cat# 2H10D10 |
| Anti-TNF-alpha Rabbit pAb | Servicebio | Cat# GB11188 |
| HuC/HuD Monoclonal Antibody | Thermo Fisher Scientific | Cat# 16A11 |
| Chemicals, peptides, and recombinant proteins | | |
| Collagenase 1A | Sigma | Cat# C2674-500mg |
| DNase I | Roche | Cat# 11284932001 |
| DMEM | Thermo Fisher Scientific | Cat# C11995500BT |
| FBS | GIBCO | Cat# A3160802 |
| OCT | Tissue-Tek | Cat# SAKURA Tissue 4583 |
| 2-Methylbutane | macklin | Cat# M875875-100ml |
| Acetone | SINOPHARM | Cat# 10000418 |
| osmic acid | Ted Pella Inc | Cat# 18456 |
| Nitroprusside disodium dihydrate | MCE | Cat# HY-A0119-50g |
| Nω-Propyl-L-arginine hydrochloride | MCE | Cat# HY-102062A-5mg |
| AGN193109 | TOCRIS | Cat# 5758 |
| A1120 | TOCRIS | Cat# 3793 |
| NCT501 | TOCRIS | Cat# 5934 |
| DAPI | Thermo Fisher Scientific | Cat# D1306 |
| Critical commercial assays | | |
| Chromium Next GEM Single Cell 5' Kit v2 | 10x Genomics | Cat# 1000263 |
| Chromium Single Cell Human BCR Amplification Kit | 10x Genomics | Cat# 1000253 |
| Masson staining kit | Servicebio | Cat# G1006-100ML |
| Mitochondrial Staining Reagent | abcam | Cat# ab176832 |
| HU Apolipoprotein (Apo) Panel (11-plex) w/ VbP | Biolegend | Cat# 50-208-8142 |
| QIAamp DNA Mini Kit | Qiagen | Cat# 51304 |
| NEBNext® Ultra™ II DNA Library Prep Kit for Illumina® | NEB | Cat# E7645L |
| Agencourt AMPure XP SPRI beads | Beckman Coulter | Cat# A63882 |
| SureSelect Human All Exon V6 Kit | Agilent | Cat# 54898 |
| NEBNext Ultra II RNA Library Prep Kit for Illumina | New England Biolabs Inc | Cat# E7775 |
| HiScript III RT SuperMix for qPCR (+gDNA wiper) | Vazyme | Cat# R323-01 |
| ChamQ Universal SYBR qPCR Master Mix | Vazyme | Cat# Q711-02 |
| FluoZin™-3 | Invitrogen | Cat# F24195 |
| AlphaTSA Multiplex IHC Kit | alphaxbio | Cat# AXT37100031 |
| Electron Microscope Fixatives | Servicebio | Cat# G1102 |
| Neurobasal™-A | Thermo Fisher Scientific | Cat# 10888022 |
| 812 embedding agent | SPI | Cat# 90529-77-4 |
| DMEM | GIBCO | Cat# 11966025 |
| Visium Spatial Tissue Optimization Slide & Reagents Kit | 10x Genomics | Cat# 1000193 |
| Visium Spatial Gene Expression Slide & Reagents Kit | 10x Genomics | Cat# 1000187 |
| Anti-Fluorescence Quenching Agent | Leagene | Cat# IH0252 |
| Annexin V-FITC/PI Apoptosis Assay kit | Invigentech | Cat# PI0100 |
| Human Retinol binding protein 4 ELISA Kit | abcam | Cat# ab108897 |
| Total nitric oxide detection kit | Beyotime | Cat# S0024 |
| Cell and tissue lysis buffer | Beyotime | Cat# S3090 |
| cGMP Assay Kit - Direct Immunoassay | abcam | Cat# ab65356-96T |
| Deposited data | | |
| Raw sequencing data | Archive of Beijing Institute of Genomics, Chinese Academy of Sciences | GSA-Human accession: HRA006505 |
| Source data | Zenodo repository | Source data is available upon reasonable request to the authors |
| Experimental models: Cell lines | | |
| Mouse embryonic fibroblasts were isolated from mouse embryos (12.5-14 d, both male and female). | This paper | NA |
| SH-SY5Y cells | Procell | Cat#CL-0208 |
| Experimental models: Organisms/strains | | |
| Sox10WT/MUT C3H mice | Jackson lab | Cat# 000290 |
| C57BL/6 mice | Charles River | Strain Code 027 |
| Oligonucleotides | | |
| *TNF -*forward: CACTTTGGAGTGATCGGCCC | This paper | NA |
| *TNF-*reverse:  GGACCTGGGAGTAGATGAGGT | This paper | NA |
| *CCL2 -*forward:  TCTGTGCCTGCTGCTCATAG | This paper | NA |
| *CCL2-*reverse:  GGGCATTGATTGCATCTGGC | This paper | NA |
| *ENTPD2 -*forward:  TCACACACGTCCATGTTTATCT | This paper | NA |
| *ENTPD2-*reverse:  AGAAGGGTTGTCTGCATAGC | This paper | NA |
| *ECRG4 -*forward:  CCTACGGCTTTAGGCATGGA | This paper | NA |
| *ECRG4-*reverse:  TTCTTGTACAGCGTGTGGCA | This paper | NA |
| *SLC30A1 -*forward:  GTTCCGTGTGAACTTGCCTG | This paper | NA |
| *SLC30A1-*reverse:  CCTTTCCAGAAGGGGCTTGT | This paper | NA |
| *APOD -*forward:  GCATCCAGGCCAACTACTCA | This paper | NA |
| *APOD-*reverse:  GGGTGGCTTCACCTTCGATT | This paper | NA |
| *CLDN1 -*forward:  CTGTCATTGGGGGTGCGATA | This paper | NA |
| *CLDN1-*reverse:  CTGGCATTGACTGGGGTCAT | This paper | NA |
| *Tnf -*forward:  CTGAGTTCTGCAAAGGGAGAG | This paper | NA |
| *Tnf-*reverse:  CCTCAGGGAAGAATCTGGAAAG | This paper | NA |
| *Ccl2 -*forward:  AGTAGGCTGGAGAGCTACAA | This paper | NA |
| *Ccl2-*reverse:  GTATGTCTGGACCCATTCCTTC | This paper | NA |
| *Sox10 -*forward:  TGGTTGCTCCAGCACTCATA | This paper | NA |
| *Sox10-*reverse:  TCTGGGTTCCCATCTGACAT | This paper | NA |
| Software and algorithms | | |
| Custom analysis code | This paper | Code is available upon reasonable request to the authors |
| ANNOVAR^7^ | <https://annovar.openbioinformatics.org/en/latest/> | 2020-06-08 |
| bwa^2^ | <https://github.com/lh3/bwa> | 0.7.17 |
| Cell Ranger | https://support.10xgenomics.com/single-cell-gene-expression/software/pipelines/latest/using/tutorial_ov | 6.1.1 |
| CellChat^13^ | <http://www.cellchat.org/> | 1.1.0 |
| ComplexHeatmap^17^ | https://github.com/jokergoo/ComplexHeatmap | 2.6.2 |
| Docker | https://www.docker.com/ | 20.10.8 |
| dplyr | https://dplyr.tidyverse.org/ | 1.0.6 |
| fastp^1^ | <https://github.com/OpenGene/fastp> | 0.23.2 |
| GATK^5^ | [https://gatk.broadinstitute.org](https://gatk.broadinstitute.org/) | 4.2.0.0 |
| ggplot2 | https://ggplot2.tidyverse.org | 3.3.6 |
| java | https://www.java.com/ | 1.8.0_262 |
| Monocle 3^11, 12^ | <https://github.com/cole-trapnell-lab/monocle3> | 1.0.0 |
| org.Hs.eg.db | https://bioconductor.org/packages/release/data/annotation/html/org.Hs.eg.db.html | 3.12.0 |
| Perl | https://www.perl.org/ | 5.26.2 |
| pheatmap | https://www.rdocumentation.org/packages/pheatmap | 1.0.12 |
| picard^4^ | <https://github.com/broadinstitute/picard> | 2.21.4 |
| Python | https://www.python.org/ | 3.8.3 |
| R | https://www.r-project.org/ | 4.0.5 |
| reshape2 | https://github.com/hadley/reshape | 1.4.4 |
| samtools^3^ | http://www.htslib.org/ | 1.13 |
| scanpy^9^ | https://scanpy.readthedocs.io/en/stable/ | 1.7.1 |
| Seurat^8^ | https://github.com/satijalab/seurat | 4.0.1 |
| SeuratDisk | https://github.com/mojaveazure/seurat-disk | 0.0.0.9019 |
| spaceranger | https://www.10xgenomics.com/support/software/space-ranger | 1.3.0 |
| STAR^18^ | https://github.com/alexdobin/STAR | 2.7.4a |
| vcf2maf | https://github.com/mskcc/vcf2maf | 1.6.21 |
| VEP^6^ | <https://useast.ensembl.org/info/docs/tools/vep> | 104 |
| Graphpad prism | https://www.graphpad.com/ | 9 |
| FlowJo | https://www.flowjo.com/ | 10.4 |
| ImageJ | https://fiji.sc/ | 2 |

**Supplementary reference**

1.	Chen S, Zhou Y, Chen Y, et al. fastp: an ultra-fast all-in-one FASTQ preprocessor. Bioinformatics 2018;34:i884-i890.

2.	Li H, Durbin R. Fast and accurate long-read alignment with Burrows-Wheeler transform. Bioinformatics 2010;26:589-95.

3.	Danecek P, Bonfield JK, Liddle J, et al. Twelve years of SAMtools and BCFtools. Gigascience 2021;10.

4.	Institute B. Picard Toolkit: GitHub Repository, 2019.

5.	McKenna A, Hanna M, Banks E, et al. The Genome Analysis Toolkit: a MapReduce framework for analyzing next-generation DNA sequencing data. Genome Res 2010;20:1297-303.

6.	McLaren W, Gil L, Hunt SE, et al. The Ensembl Variant Effect Predictor. Genome Biol 2016;17:122.

7.	Wang K, Li M, Hakonarson H. ANNOVAR: functional annotation of genetic variants from high-throughput sequencing data. Nucleic Acids Res 2010;38:e164.

8.	Hao Y, Hao S, Andersen-Nissen E, et al. Integrated analysis of multimodal single-cell data. Cell 2021;184:3573-3587 e29.

9.	Wolf FA, Angerer P, Theis FJ. SCANPY: large-scale single-cell gene expression data analysis. Genome Biol 2018;19:15.

10.	Polanski K, Young MD, Miao Z, et al. BBKNN: fast batch alignment of single cell transcriptomes. Bioinformatics 2020;36:964-965.

11.	Trapnell C, Cacchiarelli D, Grimsby J, et al. The dynamics and regulators of cell fate decisions are revealed by pseudotemporal ordering of single cells. Nat Biotechnol 2014;32:381-386.

12.	Qiu X, Mao Q, Tang Y, et al. Reversed graph embedding resolves complex single-cell trajectories. Nat Methods 2017;14:979-982.

13.	Jin S, Guerrero-Juarez CF, Zhang L, et al. Inference and analysis of cell-cell communication using CellChat. Nat Commun 2021;12:1088.

14.	Newman AM, Steen CB, Liu CL, et al. Determining cell type abundance and expression from bulk tissues with digital cytometry. Nat Biotechnol 2019;37:773-782.

15.	Reynolds G, Vegh P, Fletcher J, et al. Developmental cell programs are co-opted in inflammatory skin disease. Science 2021;371.

16.	Drokhlyansky E, Smillie CS, Van Wittenberghe N, et al. The Human and Mouse Enteric Nervous System at Single-Cell Resolution. Cell 2020;182:1606-1622 e23.

17.	Gu Z, Eils R, Schlesner M. Complex heatmaps reveal patterns and correlations in multidimensional genomic data. Bioinformatics 2016;32:2847-9.

18.	Dobin A, Davis CA, Schlesinger F, et al. STAR: ultrafast universal RNA-seq aligner. Bioinformatics 2013;29:15-21.
